# Single-cell transcriptional landscape of peripheral immunity and biomarkers for human sporadic amyotrophic lateral sclerosis patients

**DOI:** 10.1101/2022.07.03.22277129

**Authors:** Albert Ziye Ma, Brian D. Kirk, Deyu Yan, Mingchao Zhang, Jing Yao, Xiaomin Dai, Lingdan Lu, Qinghua Wang, Weihai Ying, Yan Han

## Abstract

Sporadic amyotrophic lateral sclerosis (ALS) is a fatal adult-onset neurodegenerative disease with unknown causes and few treatment options. Here we report the first multi-dimensional single-cell landscape of the peripheral blood mononuclear cell samples from seven ALS patients representing three distinct disease stages: Early, Mid and Late. Our multipronged analyses revealed the over-activation of immune cells especially in the Mid and Late ALS stages, which is characterized by the increased cytotoxic CD8^+^ effector cells, expanded TCR clonotypes, upregulated oxidative phosphorylation and ribosomal genes across many cell types. More importantly, the current study discovered novel genes strongly correlated with ALS conditions, such as *FOXO1* and *TGFBR2* responsible for the aberrant T cell activation, and *XBP1* and *SPIB* accountable for excessive oxidative stress. We further identified 60 genes that distinguished ALS patients from healthy donors with an unprecedented accuracy of 93% against an independent cohort of 792 specimens. Our findings provide new molecular and cellular insights into the mechanisms of ALS onset and progression, and offer novel strategies for accurate diagnosis and potential therapeutic development.

## Introduction

Sporadic amyotrophic lateral sclerosis (ALS) is an untreatable, adult-onset progressive neurodegenerative disease that causes motor neurons in the brain and spinal cord to die, leading to eventual respiratory muscle paralysis and death^1,2^. The average life span following diagnosis is only 2∼5 years. More than 90% of all ALS cases are sporadic without known causes, while the remaining 10% are familial ALS cases with mutations in genes such as TAR DNA-binding protein 43 (*TDP-43*), Cu-Zn superoxide dismutase (*SOD1*), heterogeneous nuclear ribonucleoprotein P2 (also known as Fused in Sarcoma, *FUS*), and *C9orf72*.

Dysfunctional peripheral immune cells contribute to motor neuron degeneration in ALS patients and model animals. Immune profiling studies of peripheral blood samples from ALS patients uncovered extensive immunophenotypic differences, including augmented T cell activation, as well as increased classical and non-classical monocytes and natural killer (NK) cells^3–7^. In addition, the numbers of regulatory T cells were found to be inversely associated with ALS progression rates, while expansion of regulatory T cells helped preserve motor neuron soma size in mice with SOD1 G93A mutant^8–10^. The transitions of peripheral immune cells such as monocytes, macrophages and regulatory T cells from anti-inflammatory to pro-inflammatory are linked to ALS pathophysiology^3,11^. Furthermore, hyperphosphorylated and ubiquitinated TDP-43-positive neuronal cytoplasmic inclusions were found in the brain and spinal cord of most ALS patients^12^. TDP-43 aggregates in neuronal cells sequestered specific microRNAs and proteins, in particular nuclear-genome-encoded mitochondrial proteins, resulting in a global mitochondrial imbalance and increased oxidative stress^13^.

Reliable peripheral blood biomarkers of ALS could enable early and accurate diagnosis, improve patient management and survival, and probably provide novel therapeutic targets. Towards this end, 14 serum proteins with significant changes in ALS patients compared to healthy controls were identified using quantitative discovery proteomics^14^. RNA-seq analysis of coding and long non-coding RNA (lncRNA) in peripheral blood mononuclear cells (PBMCs) uncovered 293 differentially expressed (DE) lncRNA and 87 mRNA between healthy donors and ALS patients^15^. Furthermore, analysis on extensive microarray data from blood samples of ALS patients (n=396) and healthy donors (n=645)^16^ identified 850 genes that distinguished ALS patients with an overall 87% accuracy (sensitivity of 86% and specificity of 87%)^17^, the highest accuracy in the literature.

Single-cell sequencing technology is a revolutionary tool that, different from bulk RNA sequencing, can better account for the high heterogeneity that ALS samples likely display. A recent study using single-cell RNA-sequencing (scRNA-seq) on the brainstems of mutant SOD1 mice discovered perturbed pathways including inflammation, stress response and mitochondrial function^18^. Most recently, a scRNA-seq study of ALS4 (caused by mutations in senataxin gene) uncovered an immunological signature consisting of clonally expanded, terminally differentiated effector memory CD8^+^ T (T_EM_) cells in the blood and central nervous system of knock-in mice^19^. The scRNA-seq of cerebrospinal fluid samples from ALS patients confirmed that cytotoxic CD4^+^ and CD8^+^ T cells were clonally expanded and the T cell profile in peripheral blood predicted ALS progression well^20^.

In the current study, by combining scRNA-seq together with single-cell T cell receptor (TCR) sequencing (scTCR-seq), we unraveled for the first time the cellular landscape of PBMC samples at single-cell resolution from sporadic ALS patients at three distinct stages: Early, Mid and Late. We further mapped out the detailed abundance changes of different PBMC cell types, discovered aberrant immune activation attributed to the downregulation of *FOXO1* and the TGFβ signaling pathway, and identified 60 genes that collectively distinguished ALS patients from healthy volunteers at 93% accuracy. Our findings provide critical new light on ALS pathogenesis and provide new biomarkers for accurate diagnosis and potential therapeutic development.

## Results

To explore the transcriptome and TCR landscape of peripheral immune cells in ALS patients, we performed scRNA-seq and scTCR-seq on PBMC cells collected from seven clinically confirmed sporadic ALS patients and five healthy donors **(Fig .1a, Fig.S1a**). The ALS Functional Rating Scale-Revised (ALSFRS-R) was utilized to score the severity of ALS patients, where lower scores indicate a more severe state of the disease. According to their ALSFRS-R scores **(Fig.S1a, Supplementary Table 1**), the seven patients were classified into three groups: Early group (n=2; average ALSFRS-R score of 46.5), Mid group (n=3; average ALSFRS-R score of 35.3), and Late group (n=2; average ALSFRS-R score of 21). PBMC cells from healthy donors served as the Control group (n=5) for comparison.

**Figure 1.**
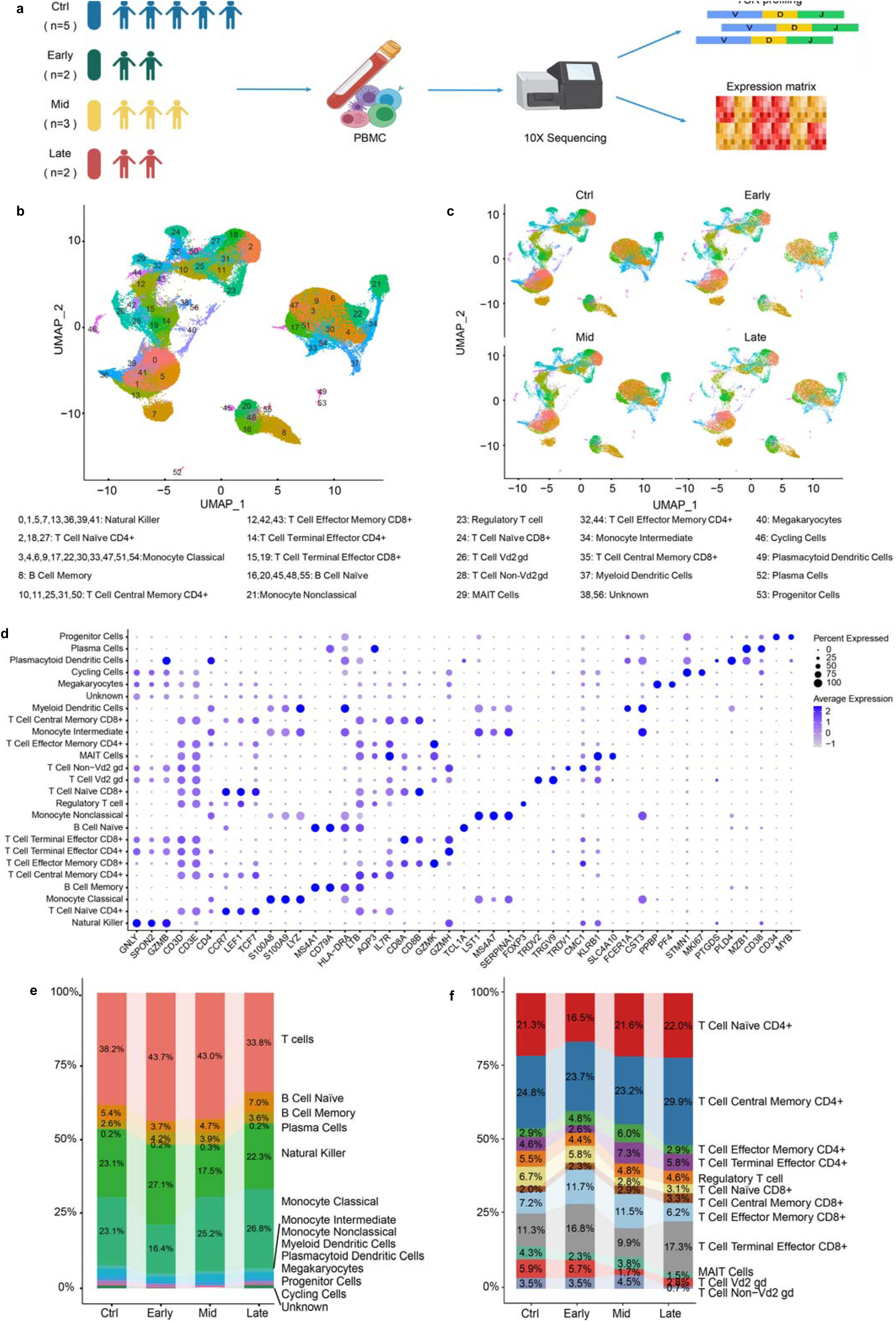
Transcriptome landscape of human PBMCs from seven ALS patients and five healthy donors. **a).** A schematic showing the overall study design. **b∼c).** UMAP visualization of cell types. Each dot represents a single cell, which is colored according to the cell type. The identity of each cell type is indicated. **b).** Pooled data. **c).** The Ctrl group and the Early, Mid and Late ALS groups, respectively. **d).** Dot plot of marker genes for distinct cell types. **e).** Cell type abundance in the four groups (Ctrl, Early, Mid and Late). The proportions of all T cells are shown as a whole. **f).** Cell type abundance of each subpopulation of T cells in the four groups (Ctrl, Early, Mid and Late).

### Single-cell transcriptome of ALS patients

After multiple quality-control steps including DoubletFinder to detect and remove doublets, we obtained high-quality scRNA-seq data for a total of 122,170 PBMC cells from the combined Control and ALS groups. Clustering analysis using SingleR^21^ detected 57 clusters (Cluster 0∼56) (**Fig. 1b∼c**). The UMAPs for the PBMC cells from the four groups showed a similar distribution (**Fig. 1c, Fig.S1b**). These clusters were annotated as 25 different cell types (**Fig. 1b∼d**) based on their marker genes (**Fig. 1d, Fig.S1c, Supplementary Table 2**).

Clusters 0, 1, 5, 7, 13, 36, 39 and 41 (high in *GNLY, SPON2* and *GZMB*) were recognized as NK cells (**Fig. 1d**). Clusters 2,18 and 27 (high in *CCR7, LEF1,* and *TCF7*) were CD4^+^ naïve T cells, while Cluster 24 was CD8^+^ naïve T cells. Clusters 3, 4, 6, 9, 17, 22, 30, 33, 47, 51 and 54 were recognized as classical monocytes, for the high levels of *S100A8, S100A9* and *LYZ*. Cluster 8 was classified as memory B cells. Clusters 10, 11, 25, 31 and 50 were designated as CD4^+^ central memory T (T_CM_) cells, while Cluster 35 was CD8^+^ T_CM_ cells. Clusters 12, 42 and 43 (high in *GZMK*) were CD8^+^ T_EM_ cells, while Clusters 32 and 44 was CD4^+^ T_EM_ cells. Clusters 15 and 19 high in *CD8* and *GZMH* were assigned as CD8^+^ terminal effector T (T_TE_) cells, while Cluster 14 was recognized as CD4^+^ T_TE_ cells. Clusters 16, 20, 45, 48 and 55 were defined as naïve B cells (high in *MS4A1, CD79A*, *HLA-DRA* and *TCL1A*). Cluster 21 (high in *LST1*, *MS4A7* and *SERPINA1*) was defined as non-classical monocytes. Cluster 23 (high in *FOXP3*) was regulatory T cells. Cluster 26 was classified as Vd2 gd T cells and Cluster 28 as non-Vd2 gd T cells. Cluster 29 was assigned as mucosal-associated invariant T (MAIT) cells. Cluster 34 was recognized as intermediate monocytes. Clusters 37 and 49 were myeloid dendritic cells and plasmacytoid dendritic cells, respectively. Cluster 40 was recognized as megakaryocytes while Cluster 46 as cycling cells (high in MKI67). Cluster 52 was assigned as plasma cells and Cluster 53 as progenitor cells. Clusters 38 and 56 was assigned as unknown cell type.

The abundance of each major cell type was compared (**Fig. 1e∼f, Fig.S1d∼e, Supplementary Table 3**). In ALS groups, there was a slight increase in the proportion of memory B cells compared to the Control group (**Fig. 1e, Fig.S1d,f)**. On the other hand, the proportion of classical monocytes decreased in the Early group compared to the Control group, but then gradually increased in the Mid and Late groups (**Fig. 1e, Fig.S1d,f)**. However, the total T cells showed an opposite trend, increased in the Early group, but gradually decreased as the disease progressed (**Fig .1e, Fig.S1d∼f)**. Among the various T cell subtypes, the proportion of regulatory T cells was lower in the three ALS groups compared to the Control (**Fig. 1f, Fig.S1e)**. Similarly, the proportion of CD8^+^ naive T cells was decreased in the three ALS groups, with a more pronounced reduction observed in the Mid and Late groups (**Fig. 1f, Fig.S1e)**. Notably, two major cytotoxic cell types, CD8^+^ T_TE_ and T_EM_ cells, exhibited significant increases in the ALS groups (**Fig. 1f, Fig.S1e)**, indicating an augmented cytotoxic activity in the disease context.

### Clonal expansion of the TCR repertoire in ALS patients

The scRNA-seq and scTCR-seq data on the same samples allowed the identification of TCR information for the same cells. Analysis of scTCR-seq data yielded a total of 42,341 clonotypes with paired TCR α- and β-chains (**Fig. 2a, Supplementary Table 4**). Among them, 94.0% (39,787 cells) were mapped to T cells identified by scRNA-seq in this study (**Supplementary Table 5**). The detected TCR for non-T cells was only 6.0%, suggesting the high specificity of scTCR-seq and confirming the accuracy of scRNA-seq-based T-cell assignment.

**Figure 2.**
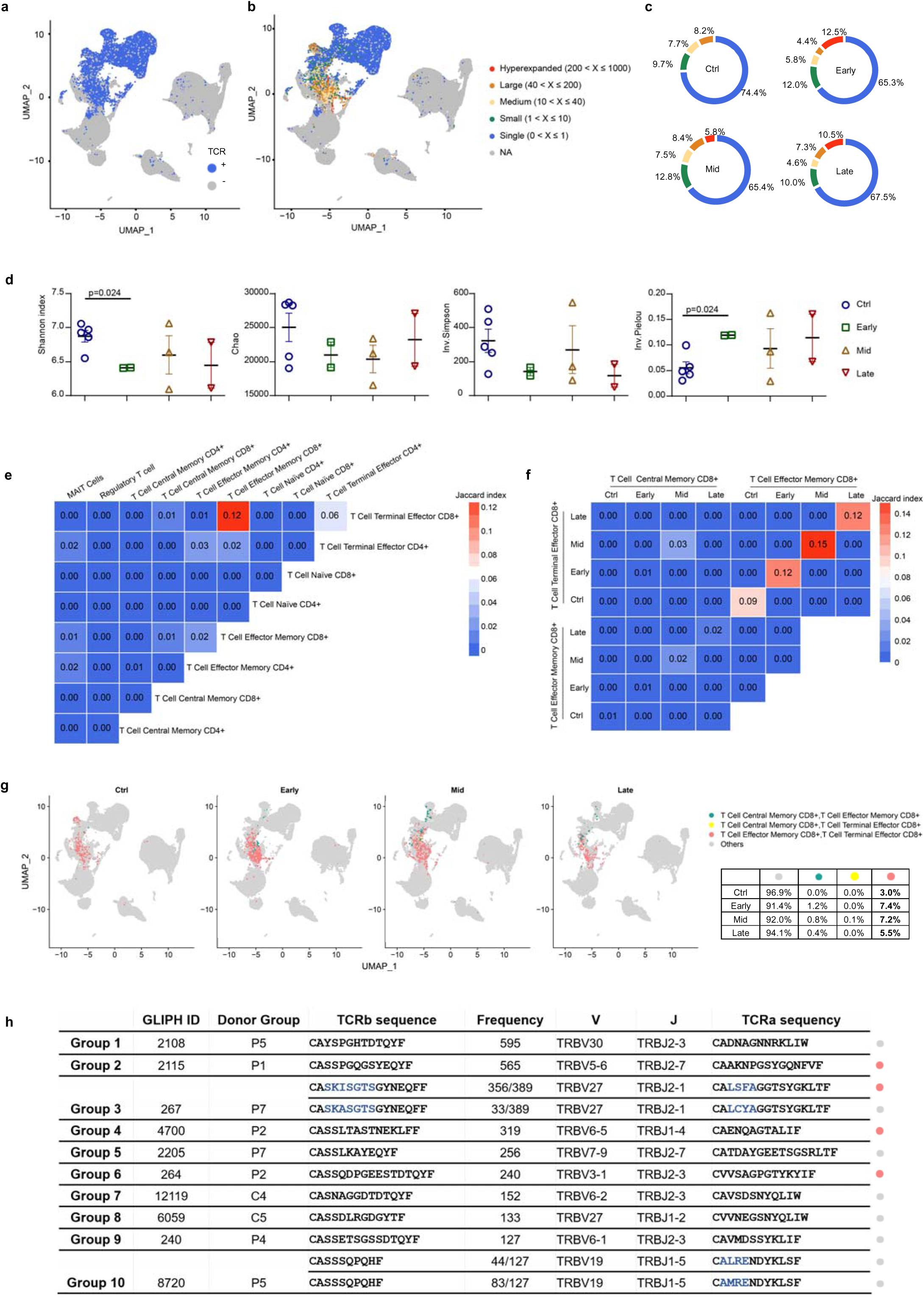
Characterization of TCR landscape. **a).** Projection of detected TCR landscape on UMAP visualization of all cells. **b).** Projection of detected TCR landscape on UMAP visualization colored according to the degrees of clonal expansion. **c).** Donut plots depicting the numbers of TCR clonotypes in each class of clonal expansion. **d).** TCR repertoire size and diversity estimation of T cells in four groups. The Shannon index (**Methods**) is a metric for quantifying clonotype diversity of the detected TCRs -higher index means higher diversity. The number of clonotypes estimated by Chao (**Methods**) indicates the predicated theoretical clonotype diversity. Inv. Simpson (**Methods**) also measures the diversity of TCR repertoire -higher score indicates higher diversity. Inv. Pielou (**Methods**) measures the evenness of TCR repertoire - lower score indicates higher evenness. **e).** The Jaccard index of shared TCR clonotypes between subsets of T cells. **f).** The Jaccard index of shared TCR clonotypes among CD8^+^ T_CM_, T_EM_ and T_TE_ cells in different groups. **g).** T cell subtype-shared TCR clonotypes projected onto UMAP visualization and colored separately. The proportion of shared TCR clonotypes to the total clonotypes is shown in the table. **h).** Top 10 Specificity TCR groups from GLIPH2 analysis with the coloring scheme as g).

Most T cell subtypes from the ALS and Control groups had a high percentage of cells with paired TCR detected (**Supplementary Table 5**). The exceptions were the non-Vd2 gd T cells (at 36% and 25% for the combined ALS group and the Control group, respectively) and Vd2 gd T cells (at 18% and 4% for the combined ALS group and the Control group, respectively). The low percentages of non-Vd2 gd T cells and Vd2 gd T cells with paired TCR α- and β-chains confirmed the correct assignment of these cell types.

The overall ratio of total TCR/unique TCR for all cell types was 1.25 for the Control group, and slightly higher, at 1.36 for the ALS groups (1.39,1.34, and 1.36 in the Early, Mid, and Late groups, respectively; **Supplementary Table 5**). As expected, TCR clonotypes in the less-differentiated T cells including CD4^+^ and CD8^+^ naïve T cells as well as CD4^+^ and CD8^+^ central memory T cells were rarely expanded (**Supplementary Table 5**). On the other hand, the more-differentiated CD4^+^ and CD8^+^ T_TE_ cells were moderately expanded, at an average ratio of 6.70 and 5.15, respectively, in the Control group, and of 7.69 and 6.67, respectively, in the ALS group (**Supplementary Table 5**). Among them, CD4^+^ and CD8^+^ T_TE_ cells in the Late ALS group were significantly highly expanded, with a ratio of 11.07 and 11.18, respectively.

We further classified TCR clonotypes by the degrees of clonal expansion in each T cell subtype: Single (0 < X≤ 1), Small (1 < X ≤ 10), Medium (10 < X ≤ 40), Large (40 < X ≤ 200), and Hyperexpanded (200 < X ≤1000) **(Fig. 2b, Fig.S2a, Supplementary Table 6)**. TCR expansion was significantly higher in the ALS groups compared to the Control group. Specifically, the portions of TCR clonotypes fell in the Hyperexpanded class were 12.5%, 5.8%, and 10.5% in the Early, Mid, and Late groups, respectively, whereas this class was absent in the Control group **(Fig .2c, Fig.S2a, Supplementary Table 6)**. The Control group exhibited a significantly higher proportion of unique TCR clonotypes (at 80.3%), while the highest proportion among the three ALS groups was observed in the Mid group, with a value of 74.8% **(Fig.S2b)**. Similarly, the percentage of unique clonotypes not shared between cell types was also of the highest in the Control group and considerably lower in all three ALS groups **(Fig.S2c)**, suggesting the more intense immune responses in ALS patients.

Next, we used several established metrics to characterize the dynamics of the TCR repertoire. The diversity was estimated using Shannon and Inverse Simpson indices, the richness bu Chao1, while the species evenness was calculated using Inverse Pielou’s index. The degree of shared TCR clonotypes between T cell subtypes was gauged using the Jaccard similarity coefficient (Jaccard index). Overall, the TCR repertoire in the Control group exhibited the highest diversities, richness and evenness, which were decreased in the three ALS groups **(Fig .2d)**. Among the different T cell subtypes, a significantly higher proportion of TCR clonotypes was found to be shared between the cytotoxic CD8^+^ T_EM_ cells and CD8^+^ T_TE_ cells, with a Jaccard coefficient of 0.12, compared to any other T cell subtypes **(Fig. 2e)**. In addition, the shared TCR clonotypes between CD8^+^ T_EM_ cells and CD8^+^ T_TE_ cells were much higher in ALS patients, with the Jaccard coefficients of 0.12, 0.15, and 0.12 for the Early, Mid, and Late groups, respectively, compared to 0.09 in the Control group **(Fig. 2f∼g, Fig.S2d)**, supporting a higher degree of T cell activation in ALS patients.

To investigate TCR specificity, we performed CDR3 pattern analysis using GLIPH2^22^ (**Supplementary Table 7**). GLIPH2 is an algorithm to cluster TCRs that recognize the same epitope according to the shared similar patterns in the CDR3 region of TCR β-chains, particularly across multiple individuals^22^. In the published study, GLIPH2 successfully identified specificity groups containing at least 3 unique TCRs from 3 or more individuals infected with *Mycobacterium tuberculosis*^22^. By using GLIPH2 on the total of 39,787 paired TCR clonotypes detected on T cells in this study, we identified 2,995 GLIPH specificity groups with at least 2 copies in each group (**Supplementary Table 7**). The top 10 most abundant specificity groups were shown in **Fig. 2h**, which had copy numbers ranging from 127∼595 copies. For the top 10 specificity groups (**Fig. 2h**), only Group 3 and Group 10 had two composite sequences. Group3 was found in P7, with the two composite sequences differing by three residues, ‘I’ *vs.* ‘A’ in the CDR3 sequences of TCR β-chains (TCRb) and ‘SF’ *vs.* ‘CY’ in the CDR3 sequences of TCR α-chains (TCRa). Group 10 was found in P5, with ‘L’ *vs.* ‘M’ in the TCRa for the two composite sequences. All other 8 GLIPH groups were composed of one single paired TCRb and TCRa sequences (**Fig. 2h**). Among these top 10 GLIPH groups, only Group 7 and 8 were from the Control group, whilst all other groups were found in the ALS group, agreeing with the higher expansion in the TCR repertoire of ALS patients (**Fig. 2c, Supplementary Table 6**). The absence of shared CDR3 patterns among different ALS donors suggests that, although ALS patients were immune-elevated, there lacked a common antigenic trigger for the immune-elevation. Among the top 10 usages, six TRBV genes (TRBV20-1, TRBV28, TRBV7-2, TRBV5-1, TRBV7-9 and TRBV19) and nine TRBJ genes (TRBJ1-1, TRBJ1-2, TRBJ1-4, TRBJ1-5, TRBJ2-1, TRBJ2-2, TRBJ2-3, TRBJ2-5 and TRBJ2-7) were shared between the ALS and Control groups (**Fig.S2e**).

### Transcriptomic signatures of PBMC cells from ALS patients

By comparing the transcriptional profiles of each cell type in the three ALS groups with the Control group, we identified 1,965, 5,871 and 5,553 DE genes for the Early, Mid, and Late groups, respectively **(Fig. 3a∼b, Supplementary Table 8∼10)**. Vd2 gd T cells in the Early group and NK cells in both Mid and Late groups had the most DE genes **(Fig. 3a)**. More DE genes were downregulated than upregulated, and the gap between them was widened as the disease progressed **(Fig. 3a∼3b)**. Among these DE genes, 307, 599 and 740 genes from the Early, Mid, and Late ALS groups, respectively, were mitochondria-encoded mitochondrial (MT) genes and ribosomal genes **(Fig. 3c)**. Furthermore, DE genes observed in 12 or more cell types out of the 24 cell types (excluding the unknown cell type) were visualized using heatmaps, which included 8, 43, and 32 for upregulated genes and 21, 55 and 52 for downregulated genes in the Early, Mid, and Late ALS groups, respectively **(Fig. 3d∼e)**.

**Fig. 3.**
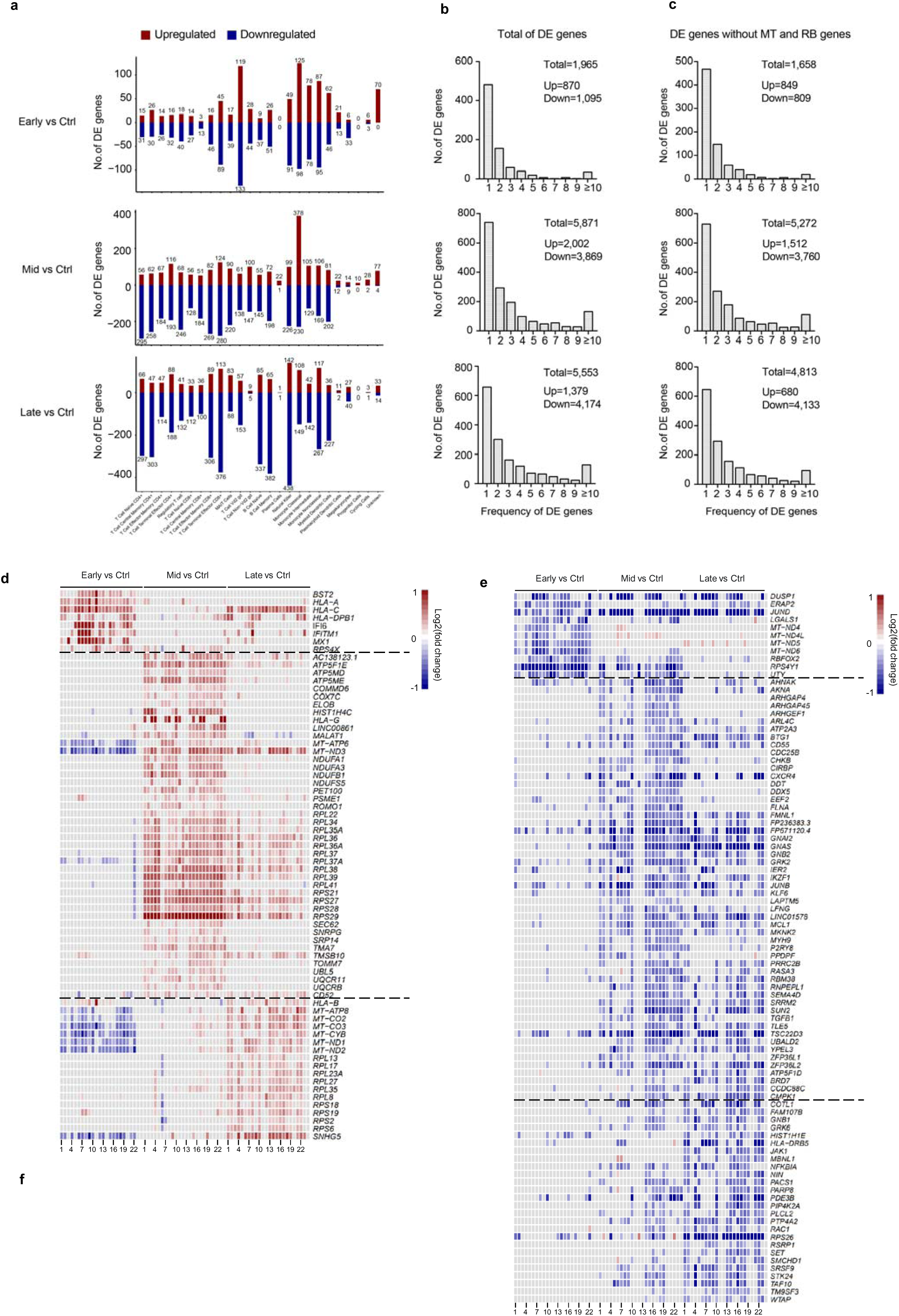
Transcriptional profiles of ALS patients. **a).** Bar charts showing the number of DE genes comparing the Control group and one of the three ALS groups. The upregulated DE genes are shown in red color while the downregulated DE genes are shown in blue color. **b).** Bar charts showing the number of DE genes between the Control group and the three ALS groups and their occurrences. **c).** Bar charts showing the number of DE genes between the Control group and the three ALS groups and their occurrences, excluding mitochondrial (MT) and ribosomal (RB) genes. **d).** Heatmap of the transcriptional levels of upregulated DE genes observed in at least twelve cell types in each ALS group. **e).** Heatmap of transcriptional levels of downregulated DE genes observed in at least twelve cell types in each ALS group. In d) and e), the order of cell types is: 1: T Cell Naïve CD4^+^; 2: T Cell Central Memory CD4^+^; 3: T Cell Effector Memory CD4^+^; 4: T Cell Terminal Effector CD4^+^; 5: Regulatory T cell; 6: T Cell Naïve CD8^+^; 7: T Cell Central Memory CD8^+^; 8: T Cell Effector Memory CD8^+^; 9: T Cell Terminal Effector CD8^+^; 10: MAIT Cells; 11: T Cell Vd2 gd; 12: T Cell Non−Vd2 gd; 13: B Cell Naïve; 14: B Cell Memory; 15: Plasma Cells; 16: Natural Killer; 17: Monocyte Classical; 18: Monocyte Intermediate; 19: Monocyte Nonclassical; 20: Myeloid Dendritic Cells; 21: Plasmacytoid Dendritic Cells; 22: Megakaryocytes; 23: Progenitor Cells; 24: Cycling Cells. **f).** Validation of *ATP5ME* and *RPS21* expression levels by qPCR. The expression levels of each gene in each ALS sample (P1∼P7) were compared to those of healthy controls (n=3), with * for *p*<0.05 and ** for *p*<0.01 in Student’s *t*-test from three independent experiments.

Interestingly, many mitochondrial and ribosomal DE genes exhibited distinct expression behaviors among the three ALS groups. For example, the majority of MT genes (such as *MT- CO2*) were upregulated in the Late group, while some nucleus-encoded mitochondrial genes (for instance *ATP5ME*) were upregulated in the Mid group **(Fig. 3d,f)**. On the other hand, some ribosomal genes (such as *RPS21*) were upregulated in the Mid group, while others including *RPL27* were upregulated in the Late group **(Fig. 3d,f)**. Due to their differential expression patterns along ALS progression stages, the mitochondrial and ribosomal DE genes were included in all subsequent analyses.

Among the upregulated genes observed in 12 or more cell types in **Fig. 3d**, there were 0 and 1 for the Early group, 2 and 14 for the Mid group, 7 and 19 for the Late group belonging to the MT or ribosomal genes, respectively. On the other hand, among the downregulated genes in Fig. 3e, 13 genes in the Early group were MT genes, while one gene in each of the Early, Mid and Late groups was ribosomal gene. Overall, these selected DE genes of high frequency were more similar to each other in the Mid and Late groups, than to those in the Early group **(Fig. 3d∼e)**.

Consistent with the more distant relationship of the Early group with the Mid and Late groups that shared a higher similarity, while the upregulated DE genes in the Early group were enriched in terms directly related to interferon signaling (e.g., *MX1* and *IFI6*) and immune effector processes (e.g., *HLA-A*, *HLA-C*, and *HLA-DBP1*) in Gene Ontology (GO) and pathway analysis, the upregulated DE genes had strong enrichment in oxidative phosphorylation (OXPHOS) (such as *MT-ND3* and *ATP5ME*) and ribosome assembly (such as *RPS29* and *RPL39*) in the Mid group, and to a lesser degree in the Late group (**Fig. 3d, Fig.S3a, Supplementary Table 11)**, indicating a sustained upregulation of mitochondrial and ribosomal activities in the later stages of disease progression.

Furthermore, while the downregulated DE genes in the Early group were predominantly enriched in the proton transmembrane transport process, and, to a lesser degree, response to oxygen levels, TGFβ signaling pathway and cell activation, the Mid group was most strongly enriched in cell activation and TGFβ signaling pathway (such as *GNAS* and *JUND*), and negative regulation of immune system process (such as *DUSP1* and *TSC22D3*), and to a slightly lesser degree for the Late group **(Fig.S3b)**. Some of these genes were significantly and consistently downregulated, exhibiting large fold changes, including the reported anti-inflammatory factor *TSC22D3* and *GNAS* (**Fig. 3e, Fig.S3c,d**). Importantly, the TGFβ signaling pathway was increasingly more strongly enriched in downregulated genes across multiple cell types going from the Early to the Mid and Late ALS groups (**Fig.S3b**), supporting a gradually aberrant immune regulation in ALS disease^23,24^.

### XBP1 and SPIB upregulate oxidative phosphorylation and ribosomal genes in ALS patients

We next performed single-cell co-expression network analysis using hdWGCNA^25^ (high dimensional weighted gene co-expression network analysis) for each cell type separately. Interestingly, we observed similar co-expression modules in different cell types that suggested an enrichment for mitochondria-related pathways **(Fig.S4a, Supplementary Table 12)**. For instance, among the 10 co-expression modules identified for the regulatory T cell subpopulation (**Fig. 4a**), the M10 module featured an enrichment for genes involved in OXPHOS (**Fig. 4b**). By evaluating the signature scores of these modules using UCell^26^, we found that in the Mid and Late groups, the M10 module had significantly higher scores compared to the Control group (**Fig.S4b**). In addition, we calculated correlation coefficients between the signature scores of M10 and ALSFRS-R scores in each ALS sample, and uncovered a strong association between the scores of M10 and ALS progression (**Fig.S4c**). Indeed, many OXPHOS genes were upregulated in one or more major cell types (T, B, NK cells and monocytes) in the ALS groups, especially the Mid and Late groups, compared to the Control group (**Fig.S4d**). Defects in energy homeostasis have been previously reported in ALS patients^27,28^. Notably, TDP-43, as the most widespread and characterized pathologic protein in ALS^29^, has been reported to bind to transcriptional messenger RNAs (mRNAs) encoding subunits of mitochondrial respiratory complex I (Complex I), leading to impaired expression and subsequent disassembly of Complex I^30,31^. In our scRNA-seq data and qPCR verification, TDP-43 expression was significantly reduced in the Mid and Late ALS groups (**Fig.S4e,f**), which may have led to a weakened inhibitory effect on the expression and assembly of Complex I, potentially contributing to the observed upregulation of Complex I in the ALS group (**Fig.S4d**). Interestingly, the observed upregulation of TDP-43 in the Early group may represent the body’s efforts to compensate some dysregulated pathways at ALS onset (**Fig.S4e,f**), agreeing with the different behaviors between the Early and the Mid/Late ALS groups (**Fig. 3, Fig.S3**).

**Fig. 4.**
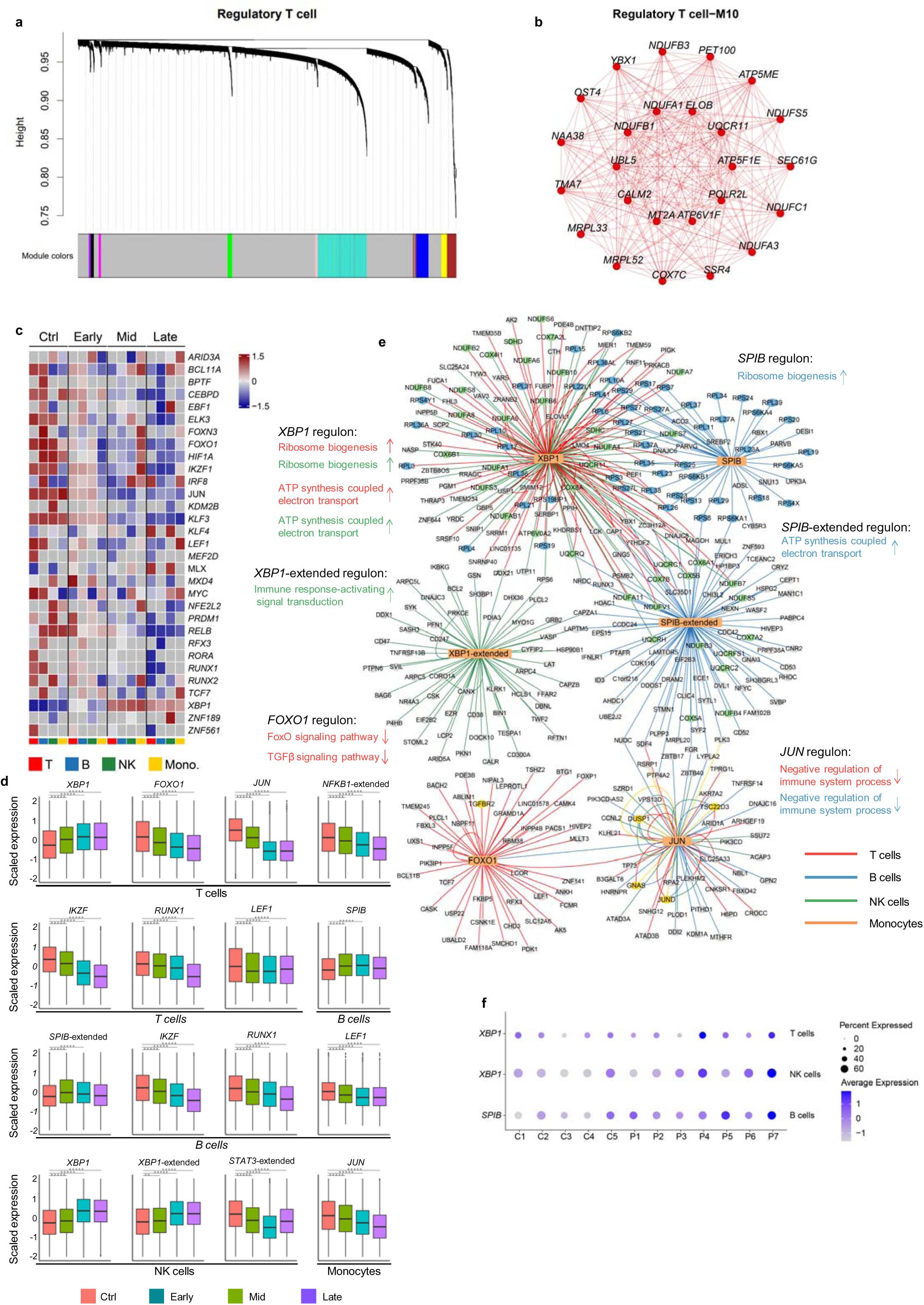
Co-expression and gene regulatory networks. **a).** Dendrogram visualization of the 10 modules of Regulatory T cell in the scale-free network. **b).** Co-expression plots for modules 10 (M10) of Regulatory T cell. **c).** Heatmap visualization of average AUC scores (regulon activities) for regulons with disease-related activity. For each group, the four columns from left to right correspond to T cells (all T cells subsets), B cells (including B cell Naïve and B cell Memory), NK cells and monocytes (including Monocyte Classical, Monocyte Nonclassical and Monocytes Intermediate). The corresponding TFs for the regulons are shown on the right of each row. The motifs of the TFs are enriched within the ±500-bp regions around the TSSs of the corresponding target genes. **d).** Box plots of mean expression levels of target genes in selected regulons. Corresponding TFs are shown above and cell types are beneath each plot. The mean expression levels were z-score transformed. ** for p<0.01 and ***** for p<10^-10^ when compared to the Control group in Wilcoxon rank-sum test. **e).** Selected regulons visualized as a network. TFs and target genes are shown as square and circular nodes, respectively. Enriched GO terms for selected regulons are shown. TF binding (shown as edges) and GO terms are color coded according to distinct cell types. ‘Extended’ indicates that the binding motifs of the TF are enriched within the ±10-kb regions around the TSSs of target genes; otherwise, the motifs are enriched in the ±500-bp regions around the TSSs. Arrows indicate the expression trend of genes in the regulons, where upwards and downwards arrows represent upregulated and downregulated expression, respectively. **e).** Dot plots of expression levels for selected TFs in distinct cell types.

To further identify key genes that broadly regulate the expression of OXPHOS genes, we performed a transcription factor (TF) regulatory network analysis using SCENIC^32^ on the four major cell types (T, B, NK cells and monocytes). By correlating expression profiles with 1,892 known human TFs, co-expression modules were identified for each TF with potential target genes. Subsequently, we performed *cis*-regulatory motif analysis to define regulons comprising TFs and target genes enriched for corresponding motifs. We then screened for regulons that showed high activity and strong negative correlation with ALSFRS-R scores, and identified 70 regulons, with 35 regulons of TFs targeting the promoter region (±500 bp around the transcription start site (TSS)) (**Fig. 4c**) and 35 regulons of TFs targeting the enhancer region (±10 kb around the TSS) (termed as TF-extended, **Fig.S4g**). Among the 35 regulons of TFs targeting the promoter regions, four TFs were observed in two cell types, including *XBP1* in T and NK cells, as well as *IKZF1, RUNX1* and *LEF1* in both T and B cells (**Fig. 4c**). Similarly, among the 35 regulons of TFs targeting the enhancer regions, four TFs were identified in more than one cell types, such as *IKZF1* in both B and NK cells, *FOXN3* in all T and NK cells and monocytes, as well as *LEF1* in both T and B cells (**Fig.S4g**). Consistent with the DE gene analyses, there were significantly more TFs with downregulated activity than those with upregulated activity in the ALS groups. Regulons targeted by the AP-1 subunit *JUN*, as well as TFs interacting with AP-1 such as *STAT3*, and the NF-κB family members *NFKB1* and *RELB*, were identified as being significantly and consistently downregulated in the ALS groups (**Fig. 4c∼d**). *FOXO1*, a TF involved in immunosuppression, was also downregulated (**Fig. 4c∼e**).

Consistent with our results from DE genes, GO and hdWGCNA analyses, a number of identified regulons involved genes in OXPHOS (**Fig. 4e)**. In T and NK cells of ALS patients, the activity of *XBP1*- regulons was significantly upregulated, with *XBP1* targeting 21 and 25 OXPHOS genes in T and NK cells, respectively (**Fig. 4c∼e)**. Furthermore, the expression of *XBP1* in these cell types was higher in ALS patients (**Fig. 4f)**. It has been reported that hypoxia in the peripheral blood is a characteristic of ALS patients^17^ and the hypoxic environment induces upregulation of *XBP1* expression^33^. Based on these findings, it is tempting to speculate that in response to hypoxic environment, *XBP1* may play a pivotal role in upregulating OXPHOS activities in T and NK cells of ALS patients. Similarly, in B cells, the *SPIB*-extended regulon upregulated 16 OXPHOS genes and the expression level of SPIB itself was also consistently upregulated in the ALS groups (**Fig. 4d∼f)**, suggesting its potential role in upregulating OXPHOS in B cells.

Noticeably, for both T cells and NK cells in ALS patients, the upregulated *XBP1*-regulon also targeted 26 ribosomal protein-coding genes in each cell type, while the *SPIB*-regulon upregulated 35 ribosomal genes in B cells (**Fig .4c∼e, Fig.S4h)**, suggesting that the OXPHOS and ribosomal coding genes were upregulated by a coupled mechanism with shared regulatory pathways in ALS patients.

### FOXO1-mediated TGF**β** pathway inhibition and immune imbalance

To explore potential alterations in ligand-receptor interactions among immune cells in ALS patients, CellChat^34^ was used to predict major signaling inputs and outputs for cells under different conditions. CellChat utilizes a comprehensive database of signaling molecule interactions and employs network analysis and pattern recognition techniques to infer intercellular communication networks. Our analysis revealed a reduction in cell-cell interactions of the TGFβ pathway in the Early ALS group and a more pronounced reduction in the Mid and Late groups compared to the Control group (**Fig. 5a∼b, Supplementary Table 13)**. Most cell types were identified as senders of TGFβ signals, with naïve T cells, monocytes, and plasma cells being among the predominant receivers (**Fig. 5b)**. As ALS progressed from the Early to Late stages, intercellular communication gradually diminished in the TGFβ signaling pathway, especially for CD8^+^ naïve T cells that exhibited a complete loss of intercellular communication as receivers in the pathway (no incoming arrows pointing to this cell) (**Fig. 5a∼b, Supplementary Table 13)**. In the predicted intercellular communication network, the TGFβ pathway was predominated by the interactions between TGFB1 (ligand) and TGFBR2 (receptor). At the gene expression level, TGFBR2 in CD8^+^ naïve T cells was significantly decreased in the more advanced Late ALS group and confirmed by qPCR analysis (**Fig.S5a∼b)**. These findings were consistent with the increasingly strong enrichment of TGFβ signaling pathway in downregulated DE genes across multiple cell types from the Early ALS group to the Mid and Late groups, including CD8^+^ naïve T cells (**Fig.S3b**).

**Fig. 5.**
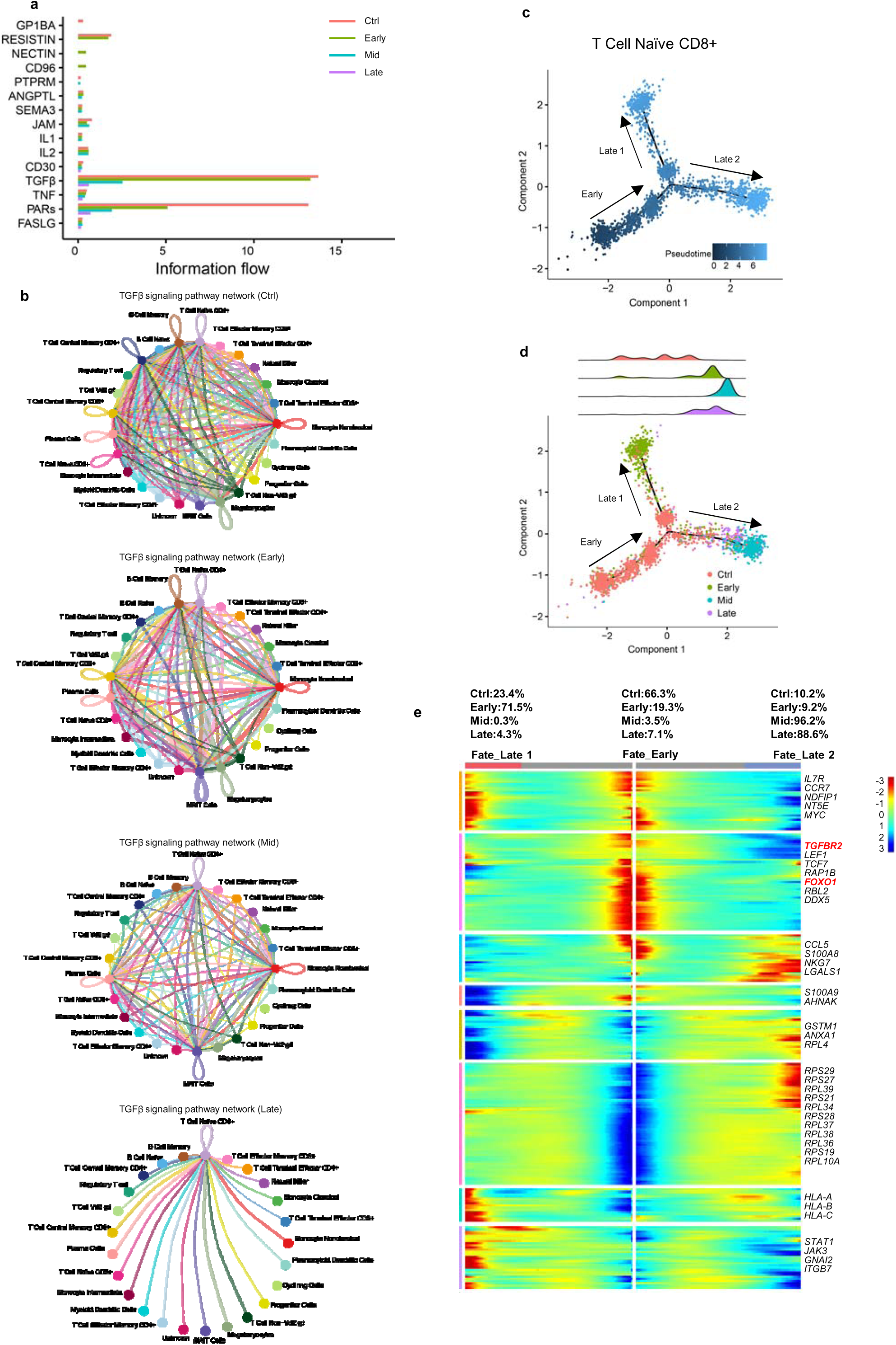
Aberrant immunity in ALS patients. **a).** Intercellular ligand-receptor interactions in the Control group and three ALS groups revealed by the overall information flow for selected signaling pathways. **b).** Chord diagrams showing the TGFβ signaling pathway for the cell-cell communication in each group. **c).** Discriminative dimensionality reduction (DDR) tree visualization of CD8^+^ naïve T cells along the pseudotime trajectory colored by pseudotime. **d).** Pseudotime analysis of CD8^+^ naïve T cells colored by groups. Top: Distribution of CD8^+^ naïve T cells from each group along the pseudotime trajectory. Bottom: DDR tree visualization of CD8^+^ naïve T cells along the pseudotime trajectory colored by groups. **e).** Heatmaps for clustering of genes that define the branchpoints (q-value, <0.05) from BEAM analysis for Early branch and Late branches (including Late 1 and Late 2) in CD8^+^ naïve T cells. Center of the gray bar above heatmap is the start of the branchpoint. The percentages of CD8^+^ naïve T cells from different groups in each branch are shown above the heatmap. Selected genes are labeled on the right with genes mentioned in the main text highlighted in red.

Previous studies demonstrated that the TGFβ signaling pathway inhibits the differentiation of CD8^+^ naïve T cells to CD8^+^ T_EM_ and T_TE_ cells^35^. In ALS patients, the diminishing level of *TGFBR2* and thereby the TGFB1-TGFBR2 interaction likely unleashed the inhibition to the differentiation of CD8^+^ naïve T cells. Indeed, there was a lower percentage of CD8^+^ naïve T cells in the Mid and Late ALS groups compared to the Control group, while the percentage of CD8^+^ effector T cells (T_EM_ and T_TE_ cells) was increased in these ALS groups (**Fig. 1f, Fig.S5c)**.

Our TF regulon analysis using SCENIC revealed that FOXO1 regulated the expression of TGFBR2 in T cells (**Fig. 4e)**. Furthermore, the expression of FOXO1 and its target regulon were significantly downregulated in T cells of ALS patients, especially in the Mid and Late groups (**Fig. 4c∼d, Fig.S5a∼b**). Combining these present findings with previous reports^36^, it is likely that the decreased expression of FOXO1 in T cells of ALS patients contributed to the downregulation of TGFBR2, leading to the weakened inhibition of T cell activation^37^ by TGFβ pathway. Importantly, FOXO1 is also a key factor on which the development of regulatory T cells depends^37^, thus the diminishing level of FOXO1 could simultaneously hamper the development of regulatory T cells, and potentially contribute to the autoimmune aspects of ALS diseases. In the ALS groups, we did observe a reduced population of regulatory T cells (**Fig.S5c)**.

Interestingly, our SCENIC analysis suggested that the transcription factor *JUN* regulated the expression of a number of genes in T cells including *FOXO1, GNAS, JUND, DUSP1* and *TSC22D3* (**Fig. 4e)**. The level of *JUN* for all cells within each sample was plotted in **Fig.S5a**, which was clearly diminished in all ALS patient samples, reminiscent of the expression profiles of *FOXO1* (**Fig.S5a∼b**), as well as of *JUND, DUSP1, GNAS* and *TSC22D3* in **Fig.S3c∼d**.

Next, we performed a pseudotime analysis on CD8^+^ naïve T cells (**Fig. 5c∼d)**. The results showed that the majority of CD8^+^ naïve T cells from the Control group (66.3%) were mapped to the Early branch, representing the beginning of the pseudo-temporal trajectory. Cells from the ALS groups were unevenly split into Late 1 and Late 2 branches. Specifically, cells from the Early ALS group (71.5%) were predominantly mapped to the Late 1 branch, while cells from the Mid and Late groups were primarily mapped to the Late 2 branch (96.2% and 88.6%, respectively). Gene expression analysis along the pseudotime trajectory revealed that genes associated with cell development and activation, such as *TCF7, CCR7* and *LEF1*, known as the hallmark genes for naïve T cells, showed high expression in the Early branch (**Fig. 5e)**. The Late 1 branch contained cells with high expression for immune effector genes such as *HLA-A, HLA-B* and *HLA-C*. In the Late 2 branch that were predominately cells from the Mid and Late ALS groups, ribosomal protein-coding genes such as *RPS21, RPS29* and *RPL39* were highly expressed (**Fig. 5e)**. Interestingly, *FOXO1* and *TGFBR2* were specifically highly expressed in the Early branch, and gradually reduced in the Late 1 branch and diminished in the Late 2 branch (**Fig. 5e)**, agreeing with the trend of the TGFβ signaling pathway shown in **Fig. 5a∼b** for the four groups.

The pseudotime analysis was also performed for regulatory T cells (**Fig.S5d∼e)**. Cells from the Control group predominantly mapped to the Early branch (69.0%). Cells from the Early ALS group were almost evenly distributed between the Early and Late 1 branches (50.9% and 46.5%, respectively), while cells from the Mid and Late ALS groups mostly fell into the Late 2 branch. Similar to CD8^+^ naïve T cells, the expression levels of *FOXO1* and *TGFBR2* were high in the Early branch and significantly decreased along the trajectory (**Fig. S5f)**. Similarly, the expression levels of *JUN*, *JUND*, *GNAS*, and *TSC22D3* were increasingly reduced from the Early, Late 1 branches to the Late 2 branch (**Fig.S5f**).

### Prediction of ALS diagnosis using gene lists derived from this study

One of the goals of this study was to seek reliable blood biomarkers of ALS to facilitate early and accurate diagnosis, thus improving patient survival. An extensive microarray analysis on blood samples of ALS (n=396) and healthy controls (n=645) identified 850 genes that distinguished ALS patients from healthy controls with 87% accuracy (sensitivity of 86% and specificity of 87%)^17^, the highest accuracy reported to date.

In order to generate a more accurate prediction model, we first identified four DE gene lists based on our current study. The first gene list (Gene List 1: Ctrl vs Others) was derived from separately comparing each cell type in the Control group and the combined ALS group (P1∼P7), and selecting genes that were differentially expressed in at least two cell types. The remaining three DE gene lists were obtained by performing the comparisons between each of the three ALS groups and the Control group separately (Gene List 2: Early vs Ctrl; Gene List 3: Mid vs Ctrl; Gene List 4: Late vs Ctrl). Subsequently, Gene List 1 was merged with each of Gene Lists 1∼3 or all of Gene Lists 1∼3 to generate Gene Lists 5∼8 as follows: Gene List 5 (Gene List 1+2), Gene List 6 (Gene List 1+3), Gene List 7 (Gene List 1+4), and Gene List 8 (Gene List 1+2+3+4) (**Supplementary Table 14**). These gene lists were used for initial feature selection towards more accurate prediction of ALS diagnosis against an independent cohort of microarray data for ALS patients (n=396) and healthy controls (n=396)^16^ (**Fig .6a**).

**Fig. 6.**
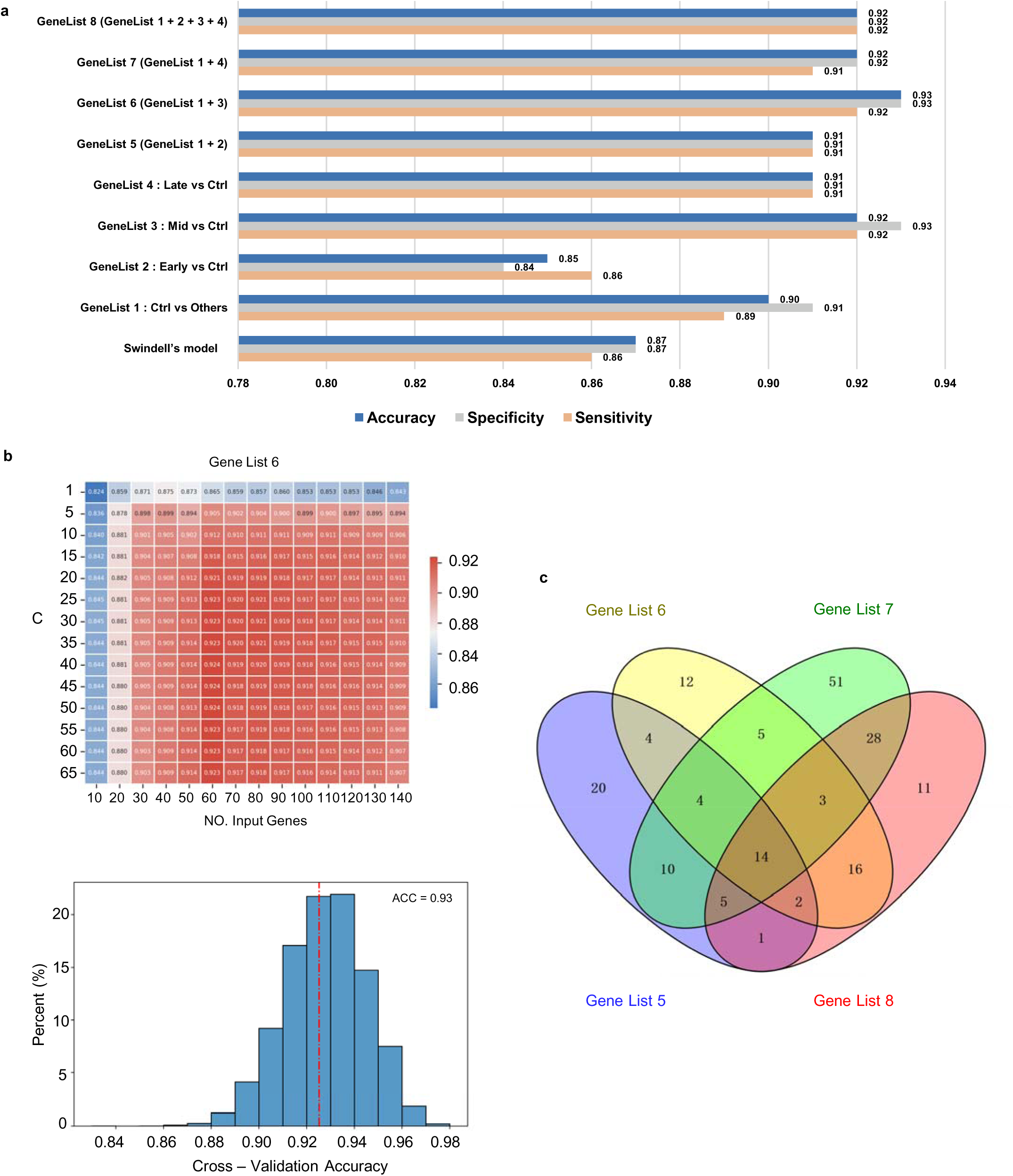
Classifiers for ALS diagnosis. **a).** Performance comparison of models using different gene lists. **b).** SVM cost and input genes parameters (top panel) and prediction accuracy (bottom panel) using Gene List 6. **c**). Venn diagram of the selected genes for Gene Lists 5∼8 used for prediction.

Starting from the total DE genes in a given Gene List, the top 300 genes with the highest correlation to label (ALS or Control) were selected after sorted by the RelieF algorithm, a feature weighting algorithm that assigns different weights to genes based on the correlation between the individual gene and the label. Subsequently the appropriate gene set was selected using the RFE algorithm. The algorithm searched for a subset of the abovementioned 300 genes, built a prediction model in the training set with the given algorithm, ranked the genes by importance and discarded the least important genes. To avoid over-fitting, the Logistic Regression algorithm was used in this process. Finally, optimal parameters were selected by grid search (**Fig. 6b**) and the model performance was evaluated using 10,000 random simulation trials (**Fig. 6c**). A total of 792 samples (396 ALS patients and 396 Control subjects)^16^ were used for model construction, of which 592 subjects were used for training (296 ALS patients and 296 Control subjects) and 200 subjects were for testing (100 ALS patients and 100 Control subjects).

Among the Gene Lists 1∼4, the predictive model based on Gene List 3 (Mid vs Ctrl) exhibited the best performance. From the 579 DE genes in Gene List 3, the average accuracy across the 10,000 simulation trials was 92% (sensitivity: 92%, specificity: 93%) using 60 input genes for ALS diagnosis (**Fig. 6a, Fig.S6a∼d, Supplementary Table 15**). The model’s performance was further enhanced after combining Gene List 3 with Gene List 1 (Control vs Others) to give rise to Gene List 6 (**Fig. 6a∼c, Supplementary Table 15**). From the 616 DE genes in Gene List 6, the average accuracy was 93% (sensitivity: 92%, specificity: 93%) with 60 genes as input. This was the best performance achieved. The constantly better performance for DE genes involving Gene List 3 (Mid vs Ctrl) could be explained by the possibility that the Mid stage is the most prevalent at diagnosis and therefore dominated the samples collected, for which the Mid group in our study is the closest proximation. To support this speculation, we randomly surveyed several publications on ALS patient studies, and found that the average ALSFRS-A scores were in the 30s^38^, agreeing with that of our Mid group.

There was a certain degree of gene overlap among the DE gene lists from feature selection (**Fig. 6d, Supplementary Table 15**). For instance, 14 genes were common among Gene Lists 5∼8 (**Fig. 6d, Supplementary Table 15**). These genes were associated with OXPHOS (such as *ATP5ME*, *NDUFA1*, and *ATP2A3*), ribosome and translation regulation (such as *RPS10*), as well as NF-κB signaling (such as *NFKB1*), suggesting that alterations in these key pathways are distinctive features of ALS pathology, agreeing with our earlier findings.

## Discussion

This study aims to shed new light on understanding the causes and improving the diagnosis of ALS. Towards this end, scRNA-seq was combined with scTCR-seq immune profiling on PBMC cells from five heathy donors and seven ALS patients. Importantly, these ALS patients represented three distinct disease stages based on their ALSFRS-A scores: Early, Mid or Late. From a total of 122,170 cells with high-quality data, we identified 25 different cell types, of them 12 cell types belonging to T cells.

Compared to the Control group, the ALS group had increased abundance in CD4^+^ and CD8^+^ T_TE_ cells and classical monocytes, as previous reported^3–6^. Moreover, the CD4^+^ and CD8^+^ T_TE_ cells from the ALS patients exhibited a higher level of clonal expansion, especially in the Late ALS group. However, out of the total of 42,341 paired TCR clonotypes detected on T cells in this study, we found no CDR3 patterns shared by more than one ALS patients. These results collectively suggest that although ALS patients exhibited a higher degree of T cell activation, there exists no common antigen responsible for T cell activation in them.

More importantly, this study uncovered the decreased abundances in CD8^+^ naïve T cells and regulatory T cells in ALS patients compared to the Control group. Pseudotime analyses revealed that these cell types in ALS patients, especially in the Mid and Late groups, exhibited a significantly different expression profiles from those in the Control group. Our multifaceted analyses support a model in which the downregulation of *FOXO1* in CD8^+^ naïve T cells of ALS patients leads to the downregulation of TGFBR2, thus disabling the TGFβ signaling pathway in CD8^+^ naïve T cells and allowing them to differentiate into cytotoxic CD8^+^ effector T cells (T_EM_ and T_TE_ cells) **(Fig. 7)**. Meanwhile, the diminished expression of *FOXO1* could hamper the normal development of regulatory T cells, accounting for the reduced population of regulatory T cells in ALS patients.

**Fig. 7.**
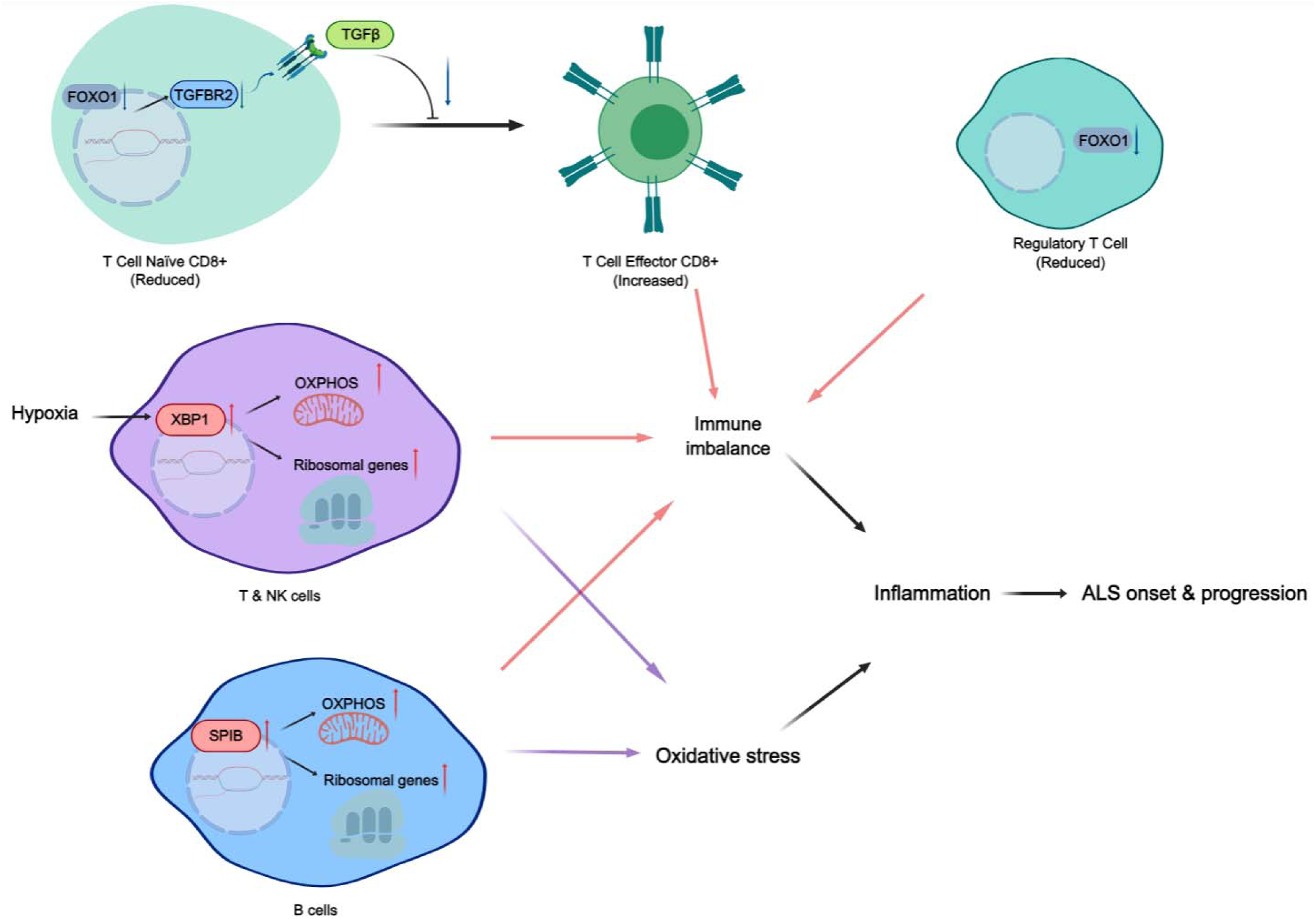
Diagram of the multifactorial ALS disease. Hypoxia-induced XBP1-mediated upregulation of OXPHOS and ribosomal genes in T and NK cells, SPIB-mediated upregulation of OXPHOS and ribosomal genes in B cells, as well as dysregulations in CD8^+^ naïve T cell differentiation and regulatory T cell development caused by FOXO1 downregulation and weakened TGFβ signaling pathway, together with other dysregulation in anti-inflammation not shown here, collectively lead to excessive oxidative stress and immune imbalance, resulting in inflammation and ALS conditions.

Additionally, multiple lines of evidence emerged from our study support a model in which *XBP1*, in response to hypoxic environment, causes higher expression levels of OXPHOS and ribosomal genes in T cells and NK cells. Similarly, SPIB induces the upregulation of a large number of OXPHOS and ribosomal genes in B cells in ALS patients. The aberrant levels of OXPHOS and ribosomal genes in these immune cells not only contribute to the augmented oxidative stress as widely noted in ALS patients^11^, but also present additional immune imbalance together with the abovementioned dysregulation of CD8^+^ naïve T cell differentiation and regulatory T cells development. The net outcome is inflammation with autoimmune characteristics, contributing to ALS onset and progression **(Fig. 7)**.

Interestingly, our TF regulon analysis using SCENIC revealed that *JUN* is responsible for the downregulation of *FOXO1* regulon in T cells, while also reducing the expression of *GNAS, JUND, DUSP1* and the anti-inflammatory factor *TSC22D3*. Furthermore, as the best characterized pathological protein in ALS, TDP-43 was believed to present toxicity to neurons by mutant proteins, protein aggregates, or various post-translational modifications^12^. For our Mid and Late ALS groups, TDP-43 had a significantly reduced expression level in scRNA-seq data as well as qPCR results, which may potentially lead to the observed upregulation of complex I in ALS patients, thereby presenting a novel mechanism in contributing to ALS progression.

Collectively, as a multifactorial disease, ALS, especially at the Mid and Late stages, exhibits multi-dimensional abnormalities from healthy donors, including dysregulated immune cells, elevated oxidative stress and impaired anti-inflammatory functions **(Fig. 7)**. The detailed mechanistic decomposition of ALS at the single-cell and molecular levels in the current study was made possible by systematic comparisons of ALS samples representing distinct progression stages: Early, Mid and Late. Given the fact that T cell profile in peripheral blood predicted ALS progression well^20^, although it remains to be deciphered how genes such as *JUN* and *TDP-43* were downregulated, these and other genes including *FOXO1, XBP1* and *SPIB* identified in this study may provide new targets for effective intervention of ALS conditions in future therapeutic investigation.

One goal of this study was to improve accuracy of ALS diagnosis. There have been a number of attempts in the literatures. Swindell *et al.*^17^ achieved the best accuracy of 0.87 using 850 genes. By using DE genes identified in comparing the three ALS groups at different disease progression stages with the Control group, here we acquired the highest prediction score of ALS diagnosis to date, with an average accuracy of 93% using 60 genes. The ALS-specific expression signatures of human peripheral blood samples from this study could constitute the foundation for formulating a clinically relevant diagnosis panel of biomarkers to enhance ALS diagnosis in clinics.

## Methods and Materials

### Blood collection and PBMC isolation

All human blood samples were collected at Yueyang Hospital of Integrated Traditional Chinese and Western Medicine, Shanghai University of Traditional Chinese Medicine (Shanghai, China), with the personal information de-identified. None of the participants had a history of prior conditions other than the clinical diagnosis of ALS for ALS donors. A total of 5 mL fresh whole blood sample was collected from each participant into a tube containing EDTA. PBMCs were isolated from whole blood within 2 hours of sample collection using Lympholyte -H (Cedarlane Labs, Cat. # C5015) following the manufacturer’s instructions.

### Single-cell transcriptome and TCR sequencing

Droplet-based encapsulation was performed using the Chromium Controller (10x Genomics). Libraries were generated using Chromium Single Cell 5’ Feature Barcode Library Kit, Chromium Next GEM Single Cell 5’ Kit, and Chromium Single Cell Human TCR Amplification Kit (10x Genomics). The libraries were multiplexed and sequenced on Illumina Novaseq 6000.

### Single-cell RNA-seq data analysis

We used the 10x Genomics Cellranger 7.0.0 pipeline^39^ to process the raw sequencing data, which included demultiplexing, read alignment (with human genome GRCh38 as the reference), barcode counting, and UMI counting. The resulting feature-barcode matrix was then imported into Seurat (v4.0.3)^40^ for downstream analysis, including quality control, normalization, dimensionality reduction, and clustering. Quality control involved first removing genes that were expressed in less than three cells across all conditions. Subsequently, cells that were identified as singlets (doublets identified by DoubletFinder (v2.0.3)^41^, had mitochondrial gene percentages <5%, ribosomal gene percentages >5%, and gene counts between 200 and 4500 were kept for downstream analysis. In order to minimize the impact of unexpected noise, genes related to mitochondria, heat-shock proteins, ribosomes, dissociation, and T-cell receptors (TCRs) were excluded prior to the clustering and cellular annotation steps. The gene expression data for each sample was normalized using the SCTransform function, which were then consolidated and integrated into a master Seurat object (IntegrateData). This master Seurat object contained 25,208 genes across 122,170 cells. The top 45 principal components (RunPCA) were used to perform dimensionality reduction on the integrated matrix via UMAP projection (RunUMAP). Various clustering resolutions were then interrogated with the clustree (v0.4.4) R package^42^, and subsequently, clustering was performed with FindClusters at a resolution of 2, yielding 57 clusters. Finally, visualization was done with the DimPlot feature.

Cell type annotations were determined with the R package SingleR (v1.4.1)^21^, which compared the transcriptome of each individual cell to three reference databases: Database of Immune Cell Expression, Novershtern hemopoietic data, and Monaco immune data. A score heatmap and delta distribution plot were produced to validate SingleR’s conclusions. SingleR annotations were then refined, returned to Seurat, and transferred to the cell clusters. The expression of selected characteristic genes was also plotted (VlnPlot) to further validate the final annotations.

### Differential expression analysis

In the differential expression analysis and subsequent analyses, the full set of genes (25,589 genes) was utilized without excluding any genes associated with mitochondria, ribosomes, or any other categories. We utilized Seurat to generate four distinct sets of DE genes. The first set of DE genes was generated with the FindAllMarkers (log_2_FC threshold of 0.25) function by extracting each cell type from all samples (for example, Monocyte Classical) and comparing its expression profile against the expression profile of all other cell types combined. DE genes with a Bonferroni-adjusted *p*-value <0.05 and an absolute log_2_FC threshold greater than 0.25 were considered significant.

The other three DE gene sets were generated by comparing the expression levels of each cell type in each ALS group with the expression levels of the same cell types in the Control group using FindMarkers. This was repeated for all 25 cell types. DE genes with absolute log_2_ fold change values greater than 0.25 and Bonferroni-adjusted *p*-values less than 0.05 were considered significant and kept for further investigation. For each group, the preserved DE genes were then passed on for GO analysis. Additionally, the expression patterns of DE genes found in twelve or more cell types were also visualized with the pheatmap (v1.0.12) R package (RRID:SCR_016418). Selected DE genes underwent further visualization with VlnPlots.

### Pseudotemporal analysis

We used the R package Monocle (v2.18.0)^43^ to perform pseudotime and branched chain analysis on T Cell Naïve CD8+ and Regulatory T cell, respectively. For each cell type, DE genes from each condition compared to the other three groups (e.g., Control vs Others; Early vs Others) were used as ordering genes for the DDRtree analysis (reduceDimension). The cells were then ordered along pseudotime with the function “OrderCells” and the cell fate trajectories were plotted. We utilized Monocle’s branched expression analysis modelling (BEAM) in order to identify the most branch-dependent genes. The top 250 most branch-dependent genes for each comparison (as determined by q-values) were then smoothed and visualized with heatmaps. The distribution of cells on each branch was shown for each sample.

### TCR repertoire analysis

Raw scTCR-seq data for each sample was assembled with the 10x Genomics Cellranger V(D)J 7.0.0 pipeline, using human reference dataset GRCh38. Following the assembly, the R package scRepertoire^44^ (v1.7.2) was used to assign clonotype based on two TCR chains and analyze the clonotype dynamics. Only TCRs with paired chains (TRA and TRB) were considered productive and preserved. The preserved data were then passed to Seurat, thus removing TCR expressing cells that did not belong to the final 122,170 cells that survived the quality control in scRNA-seq data analysis. Immune expression was projected onto the Seurat UMAP with the DimPlot function. Subsequently, clonotype abundance data for TCRs among T cell subtypes were exported for each experimental condition and visualized for both total and unique counts. The diversity of the TCR repertoire was gauged using Shannon and inverse Simpson indices. The richness of the repertoire was evaluated using Chao1, while species evenness was calculated using inverse Pielou’s index. The shared TCR clonotypes between T cell subtypes was characterized using the Jaccard similarity coefficient. The mathematical definitions of each index are as follows.

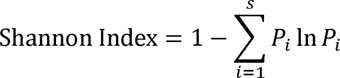

where *P_i_* represents the relative frequency of TCR clonotype *i* .

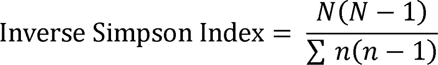

where *N* represents the total number of TCR clonotypes and *n* refers to the number of each TCR clonotype subtypes.

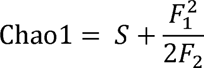

where *S* denotes the total number of unique TCR clonotypes, F_1_ refers to the count of TCR clonotypes that are observed only once (counted as 1), while F_2_ represents the count of TCR clonotypes that are observed twice (counted as 2).

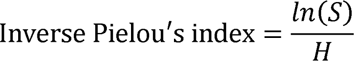

where *S* denotes the total number of unique TCR clonotypes and *H* refers to Shannon index.

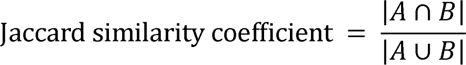

where *A* and *B* represent the TCR clonotypes of the two cell types, respectively.

CDR3β, TRBV, TRBJ, CDR3α, sample and frequency data were also passed to the GLIPH2 algorithm^22^ to test for TCR antigen specificity. Finally, V-J junction frequency for all TCR expressing cells was analyzed with VDJtools (v1.2.1)^45^. The data for the Control group and the three ALS groups were analyzed separately.

### Gene Ontology analysis

DE gene sets were passed to Metascape^46^ for GO enrichment analysis. The three DE gene sets were generated by comparing each cell type in the three ALS groups with the corresponding cell type in the Control group. Separate GO analyses for upregulated and downregulated genes were performed for each cell type within each of the three gene sets, resulting in a total of 150 runs. The most significantly enriched pathways generated from each gene list (based on absolute -log_10_*P* values) were then consolidated and visualized with dot plots in the ggplot2 (v3.3.5) R package^47^.

### Single-cell consensus weighted gene co-expression network analysis

hdWGCNA^25^ was used for co-expression network analysis on single-cell data. Each cell type was independently analyzed. Due to the hdWGCNA method’s sensitivity to sparse matrices, only genes expressed in at least 5% of the cells in the dataset were retained. The k-Nearest Neighbors (KNN) algorithm was applied to identify groups of similar cells, which were then aggregated. The average or summed expression of these cells was computed, resulting in a metacell gene expression matrix. For determining the appropriate soft power threshold in scale-free graphs, the “TestSoftPowers” function simulated co-expression networks at various thresholds. The “PlotSoftPowers” function was used to visualize the results of this parameter sweep, enabling the selection of the recommended soft threshold.

The “ConstructNetwork” function was applied to construct gene co-expression networks. During this process, a Topological Overlap Matrix (TOM) was constructed based on Pearson correlation to reflect the similarity of gene expression. The clustering tree was then plotted using the “PlotDendrogram” function. The eigengene-based connectivity, also known as kME, was calculated to assess those genes that were highly connected within each module. To gauge the significance of hub genes, we calculated the hub gene significance score using the Ucell method with the ‘ModuleExprScore’ function. Additionally, we assessed the correlation with the disease by employing the “ModuleTraitCorrelation” function. The “ModuleNetworkPlot” function was used to visualize the top 20 kME-ranked genes in the selected module.

### Gene regulatory network

The SCENIC pipeline was used^32^ (pySCENIC, v.0.11.2) to assess gene regulatory networks associated with TFs at single-cell level. The input consisted of the gene expression matrices and a list of 1,892 known human TFs (https://scenic.aertslab.org/). The GRNBoost2^48^ algorithm was used to construct matrices of co-expressed genes, relying on a tree-based regression model that is based on gradient boosting machine regression. Subsequently, the cirTarget was employed for *cis*-regulatory motif discovery of putative target genes within each co-expression module. The focus was on identifying modules that exhibited significant motif enrichment within ±500bp or ±10kb regions around the TSS of the corresponding TF. Finally, the activity of the regulons (*i.e.,* TFs and their target genes) was analyzed using the AUCell.

The SCENIC pipeline was applied to four major cell types: T cells, B cells, NK cells, and monocytes, to identify regulons that were specific to each cell type. By calculating the Pearson correlation coefficient between the activity of each TF and the ALS score of the samples (with the Control samples assigned a score of 60), we identified TFs highly negatively correlated with the disease with average area under the curve (AUC) score > 0.1 and correlation *p*-value < 0.05 for at least one cell type. Thirty-five highly correlated regulons were identified within the ±10-kb and ±500-bp regions, respectively. Heatmaps were generated to visualize the activity of these regulons under the four conditions.

### Cell–cell communication analysis

CellChat^34^ was employed to infer cell-cell interactions based on the expression of known ligand-receptor pairs in different cell types. Potential cell-cell communication networks were identified for each of the four groups (Control, Early, Mid, and Late). The normalized counts were loaded into CellChat, and preprocessing functions like “identifyOverExpressedGenes” and “identifyOverExpressedInteractions” were applied with standard parameters. CellChat was based on a comprehensive database including secreted signaling, ECM-receptor, and Cell-Cell contact relationships. For the main analyses, core functions such as computeCommunProb, computeCommunProbPathway, and aggregateNet were applied using standard parameters.

### Feature selection and machine learning construction for accurate ALS diagnosis

First, we generated four DE gene lists. This included comparing all cell types in the Control group with the overall ALS groups (Gene List 1: Ctrl vs Others), as well as conducting separate comparisons of the three ALS subgroups with the Control group (Gene List 2: Early vs Ctrl; Gene List 3: Mid vs Ctrl; Gene List 4: Late vs Ctrl). For each comparison, genes that exhibited differential expression in two or more cell types simultaneously were selected. Subsequently, Gene List 1 was combined with the other three gene lists to create an additional four gene lists (Gene List 5: Gene List 1+2; Gene List 6: Gene List 1+3; Gene List 7: Gene List 1+4; Gene List 8: Gene List 1+2+3+4). These eight gene lists were utilized as input features for the ALS diagnostic model.

We used published microarray data from blood samples of ALS patients and healthy controls (GSE112676 and GSE112680) as an independent cohort. Expression profiles were preprocessed using background correction, quantile normalization, and batch correction. For subsequent comparisons, the data processing steps were kept consistent with Swindell *et al*^17^. A total of 792 samples (296 ALS patients vs. 296 Control subjects) were used for model construction, of which 592 subjects were used for training (296 ALS patients vs. 296 Control subjects) and 200 subjects were used for testing (100 ALS patients vs. 100 Control subjects).

The importance of each gene for a given gene list was scored using the Relief (Python package: skrebate; function: ReliefF), a feature weighting algorithm that assigns different weights to features based on the relevance of each feature and category. The Top 300 ranked genes were selected for further feature combination. A fixed number of genes from the 300 mentioned above were selected using the REF algorithm (Recursive Feature Elimination, Python package: sklearn; function: RFE) and used as features for model training. Average accuracy, specificity and sensitivity were used to assess the performance of the model.

### RNA isolation, cDNA synthesis and real-time qPCR

Total RNA from PBMCs was isolated using Quick-RNA Whole Blood kit (Zymo Research) according to the manufacturer’s instruction. cDNA was synthesized using the iScript Reverse Transcription Supermix (Bio-rad) according to the manufacturer’s instruction. Real-time qPCR was performed using SYBR Green qPCR Master Mix (Bimake) on QuantStudio 6. The fold change in the expression of target genes in the ALS group compared with the Control group was calculated as 2(^−ΔΔCT^) using GAPDH as a reference gene. The sequences for qPCR primer are provided in **Supplementary Table 16**.

### Statistics

The R package was used for statistical analysis.

### Study approval

This study was approved by the Ethics Committee of the Yueyang Hospital of Integrated Traditional Chinese and Western Medicine, Shanghai University of Traditional Chinese Medicine (Approval #2020-060-02). Written informed consent was received prior to participation.

## Author contributions

A.Z.M. conceived the study, A.Z.M. and W.Y. designed the study, L.L and Y.H. provided critical clinical samples, A.Z.M. and B.D.K. together with D.Y., M.Z., J.Y. and X.D. conducted the study, A.Z.M., Q.W. and W.Y. wrote the manuscript.

## Data Availability

All data produced in the present study are available upon reasonable request to the authors

## Acknowledgement

This work was partially supported by Major Special Program Grant of Shanghai Municipality (Award Number 2017SHZDZX01), National Key Research and Development Program of China (Award Number 2021YFF1200400), Shanghai Municipal Science and Technology Major Project (Award Number 2018SHZDZX01) and PJLab.

## References

1 Beers, D. R. & Appel, S. H. Immune dysregulation in amyotrophic lateral sclerosis: mechanisms and emerging therapies. Lancet Neurol 18, 211–220 (2019). 10.1016/S1474-4422(18)30394-6

2 Mead, R. J., Shan, N., Reiser, H. J., Marshall, F. & Shaw, P. J. Amyotrophic lateral sclerosis: a neurodegenerative disorder poised for successful therapeutic translation. Nat Rev Drug Discov 22, 185–212 (2023). 10.1038/s41573-022-00612-2

3 Jin, M., Gunther, R., Akgun, K., Hermann, A. & Ziemssen, T. Peripheral proinflammatory Th1/Th17 immune cell shift is linked to disease severity in amyotrophic lateral sclerosis. Sci Rep 10, 5941 (2020). 10.1038/s41598-020-62756-8

4 Xie, Y., Luo, X., He, H. & Tang, M. Novel Insight Into the Role of Immune Dysregulation in Amyotrophic Lateral Sclerosis Based on Bioinformatic Analysis. Front Neurosci 15, 657465 (2021). 10.3389/fnins.2021.657465

5 Rolfes, L. et al. Amyotrophic lateral sclerosis patients show increased peripheral and intrathecal T-cell activation. Brain Commun 3, fcab157 (2021). 10.1093/braincomms/fcab157

6 Gustafson, M. P. et al. Comprehensive immune profiling reveals substantial immune system alterations in a subset of patients with amyotrophic lateral sclerosis. PLoS One 12, e0182002 (2017). 10.1371/journal.pone.0182002

7 Garofalo, S. et al. Natural killer cells modulate motor neuron-immune cell cross talk in models of Amyotrophic Lateral Sclerosis. Nat Commun 11, 1773 (2020). 10.1038/s41467-020-15644-8

8 Beers, D. R. et al. ALS patients’ regulatory T lymphocytes are dysfunctional, and correlate with disease progression rate and severity. JCI Insight 2, e89530 (2017). 10.1172/jci.insight.89530

9 Sheean, R. K. et al. Association of Regulatory T-Cell Expansion With Progression of Amyotrophic Lateral Sclerosis: A Study of Humans and a Transgenic Mouse Model. JAMA Neurol 75, 681–689 (2018). 10.1001/jamaneurol.2018.0035

10 Beers, D. R. et al. Endogenous regulatory T lymphocytes ameliorate amyotrophic lateral sclerosis in mice and correlate with disease progression in patients with amyotrophic lateral sclerosis. Brain 134, 1293–1314 (2011). 10.1093/brain/awr074

11 Appel, S. H. Oxidative stress-mediated inflammation promotes the pathogenesis of amyotrophic lateral sclerosis. Ageing Neur Dis 2 (2022). 10.20517/and.2022.26

12 Tamaki, Y. & Urushitani, M. Molecular Dissection of TDP-43 as a Leading Cause of ALS/FTLD. Int J Mol Sci 23 (2022). 10.3390/ijms232012508

13 Zuo, X. et al. TDP-43 aggregation induced by oxidative stress causes global mitochondrial imbalance in ALS. Nat Struct Mol Biol 28, 132–142 (2021). 10.1038/s41594-020-00537-7

14 Katzeff, J. S. et al. Altered serum protein levels in frontotemporal dementia and amyotrophic lateral sclerosis indicate calcium and immunity dysregulation. Sci Rep 10, 13741 (2020). 10.1038/s41598-020-70687-7

15 Gagliardi, S. et al. Long non-coding and coding RNAs characterization in Peripheral Blood Mononuclear Cells and Spinal Cord from Amyotrophic Lateral Sclerosis patients. Sci Rep 8, 2378 (2018). 10.1038/s41598-018-20679-5

16 van Rheenen, W. et al. Whole blood transcriptome analysis in amyotrophic lateral sclerosis: A biomarker study. PLoS One 13, e0198874 (2018). 10.1371/journal.pone.0198874

17 Swindell, W. R., Kruse, C. P. S., List, E. O., Berryman, D. E. & Kopchick, J. J. ALS blood expression profiling identifies new biomarkers, patient subgroups, and evidence for neutrophilia and hypoxia. J Transl Med 17, 170 (2019). 10.1186/s12967-019-1909-0

18 Liu, W. et al. Single-cell RNA-seq analysis of the brainstem of mutant SOD1 mice reveals perturbed cell types and pathways of amyotrophic lateral sclerosis. Neurobiol Dis 141, 104877 (2020). 10.1016/j.nbd.2020.104877

19 Campisi, L. et al. Clonally expanded CD8 T cells characterize amyotrophic lateral sclerosis-4. Nature 606, 945–952 (2022). 10.1038/s41586-022-04844-5

20 Yazdani, S. et al. T cell responses at diagnosis of amyotrophic lateral sclerosis predict disease progression. Nat Commun 13, 6733 (2022). 10.1038/s41467-022-34526-9

21 Aran, D. et al. Reference-based analysis of lung single-cell sequencing reveals a transitional profibrotic macrophage. Nat Immunol 20, 163–172 (2019). 10.1038/s41590-018-0276-y

22 Huang, H., Wang, C., Rubelt, F., Scriba, T. J. & Davis, M. M. Analyzing the Mycobacterium tuberculosis immune response by T-cell receptor clustering with GLIPH2 and genome-wide antigen screening. Nat Biotechnol 38, 1194–1202 (2020). 10.1038/s41587-020-0505-4

23 Batlle, E. & Massague, J. Transforming Growth Factor-beta Signaling in Immunity and Cancer. Immunity 50, 924–940 (2019). 10.1016/j.immuni.2019.03.024

24 Travis, M. A. & Sheppard, D. TGF-beta activation and function in immunity. Annu Rev Immunol 32, 51–82 (2014). 10.1146/annurev-immunol-032713-120257

25 Morabito, S., Reese, F., Rahimzadeh, N., Miyoshi, E. & Swarup, V. hdWGCNA identifies co-expression networks in high-dimensional transcriptomics data. Cell Rep Methods 3, 100498 (2023). 10.1016/j.crmeth.2023.100498

26 Andreatta, M. & Carmona, S. J. UCell: Robust and scalable single-cell gene signature scoring. Comput Struct Biotechnol J 19, 3796–3798 (2021). 10.1016/j.csbj.2021.06.043

27 Dupuis, L., Pradat, P. F., Ludolph, A. C. & Loeffler, J. P. Energy metabolism in amyotrophic lateral sclerosis. Lancet Neurol 10, 75–82 (2011). 10.1016/S1474-4422(10)70224-6

28 Ravera, S. et al. Characterization of the Mitochondrial Aerobic Metabolism in the Pre- and Perisynaptic Districts of the SOD1(G93A) Mouse Model of Amyotrophic Lateral Sclerosis. Mol Neurobiol 55, 9220–9233 (2018). 10.1007/s12035-018-1059-z

29 Kim, G., Gautier, O., Tassoni-Tsuchida, E., Ma, X. R. & Gitler, A. D. ALS Genetics: Gains, Losses, and Implications for Future Therapies. Neuron 108, 822–842 (2020). 10.1016/j.neuron.2020.08.022

30 Kodavati, M., Wang, H. & Hegde, M. L. Altered Mitochondrial Dynamics in Motor Neuron Disease: An Emerging Perspective. Cells 9 (2020). 10.3390/cells9041065

31 Wang, W. et al. The inhibition of TDP-43 mitochondrial localization blocks its neuronal toxicity. Nat Med 22, 869–878 (2016). 10.1038/nm.4130

32 Aibar, S. et al. SCENIC: single-cell regulatory network inference and clustering. Nat Methods 14, 1083–1086 (2017). 10.1038/nmeth.4463

33 Romero-Ramirez, L. et al. XBP1 is essential for survival under hypoxic conditions and is required for tumor growth. Cancer Res 64, 5943–5947 (2004). 10.1158/0008-5472.CAN-04-1606

34 Jin, S. et al. Inference and analysis of cell-cell communication using CellChat. Nat Commun 12, 1088 (2021). 10.1038/s41467-021-21246-9

35 Rubtsov, Y. P. & Rudensky, A. Y. TGFbeta signalling in control of T-cell-mediated self-reactivity. Nat Rev Immunol 7, 443–453 (2007). 10.1038/nri2095

36 Shi, L. et al. FoxO1 regulates adipose transdifferentiation and iron influx by mediating Tgfbeta1 signaling pathway. Redox Biol 63, 102727 (2023). 10.1016/j.redox.2023.102727

37 Kerdiles, Y. M. et al. Foxo transcription factors control regulatory T cell development and function. Immunity 33, 890–904 (2010). 10.1016/j.immuni.2010.12.002

38 Murdock, B. J., Goutman, S. A., Boss, J., Kim, S. & Feldman, E. L. Amyotrophic Lateral Sclerosis Survival Associates With Neutrophils in a Sex-specific Manner. Neurol Neuroimmunol Neuroinflamm 8 (2021). 10.1212/NXI.0000000000000953

39 Zheng, G. X. et al. Massively parallel digital transcriptional profiling of single cells. Nat Commun 8, 14049 (2017). 10.1038/ncomms14049

40 Hao, Y. et al. Integrated analysis of multimodal single-cell data. Cell 184, 3573–3587 e3529 (2021). 10.1016/j.cell.2021.04.048

41 McGinnis, C. S., Murrow, L. M. & Gartner, Z. J. DoubletFinder: Doublet Detection in Single-Cell RNA Sequencing Data Using Artificial Nearest Neighbors. Cell Syst 8, 329–337 e324 (2019). 10.1016/j.cels.2019.03.003

42 Zappia, L. & Oshlack, A. Clustering trees: a visualization for evaluating clusterings at multiple resolutions. Gigascience 7 (2018). 10.1093/gigascience/giy083

43 Qiu, X. et al. Single-cell mRNA quantification and differential analysis with Census. Nat Methods 14, 309–315 (2017). 10.1038/nmeth.4150

44 Borcherding, N., Bormann, N. L. & Kraus, G. scRepertoire: An R-based toolkit for single-cell immune receptor analysis. F1000Res 9, 47 (2020). 10.12688/f1000research.22139.2

45 Shugay, M. et al. VDJtools: Unifying Post-analysis of T Cell Receptor Repertoires. PLoS Comput Biol 11, e1004503 (2015). 10.1371/journal.pcbi.1004503

46 Zhou, Y. et al. Metascape provides a biologist-oriented resource for the analysis of systems-level datasets. Nat Commun 10, 1523 (2019). 10.1038/s41467-019-09234-6

47 Wickham, H. ggplot2: Elegant Graphics for Data Analysis. (Springer-Verlag New York, 2016).

48 Moerman, T. et al. GRNBoost2 and Arboreto: efficient and scalable inference of gene regulatory networks. Bioinformatics 35, 2159–2161 (2019). 10.1093/bioinformatics/bty916

