## Supplementary Figures S1-6 for "Single-cell transcriptional landscape of peripheral immunity and biomarkers for human sporadic amyotrophic lateral sclerosis patients"

### Slide 1
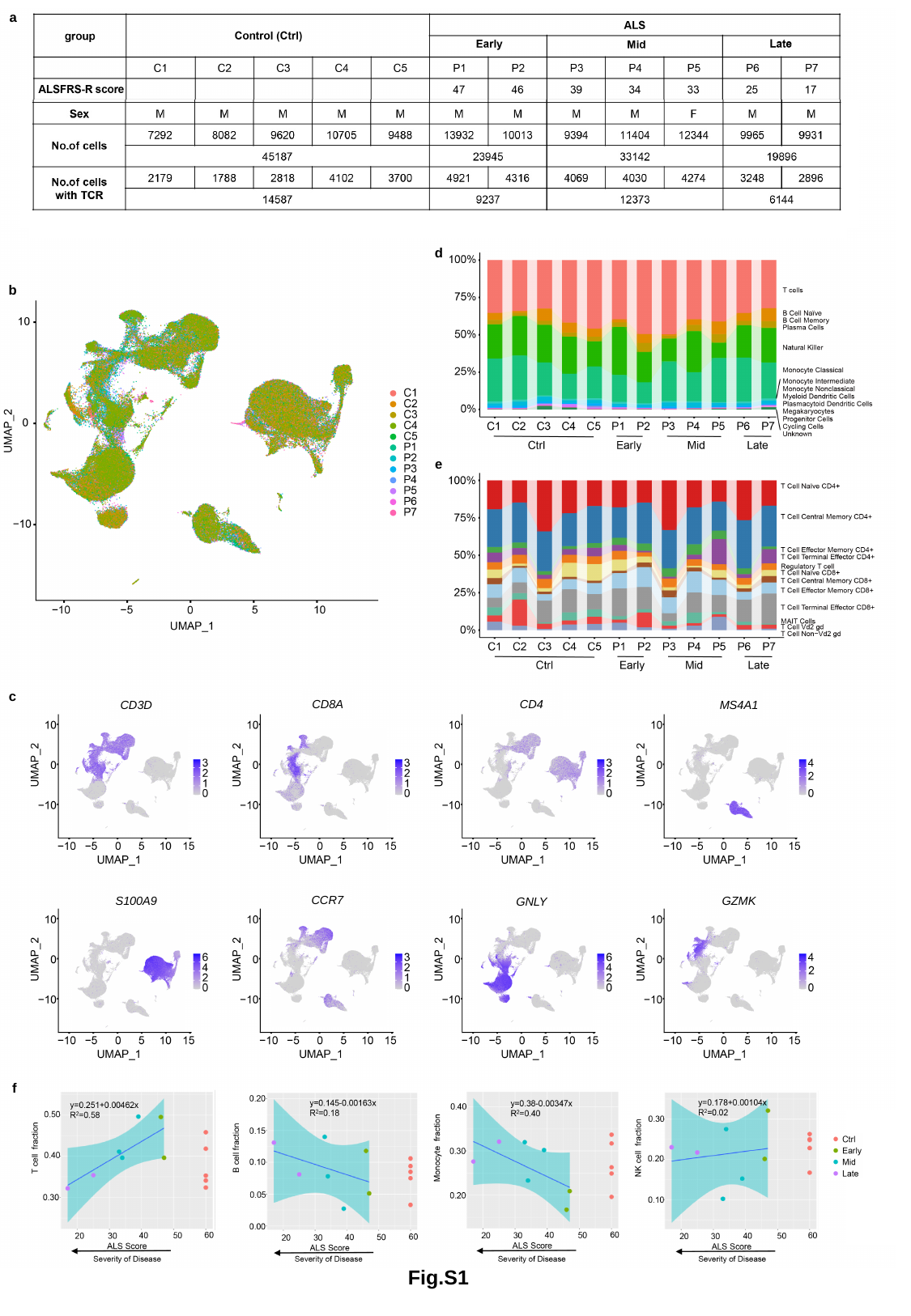

a
d
b
e
c
CD8A
CD4
CD3D
MS4A1
CCR7
S100A9
GZMK
GNLY
f
Fig.S1

### Slide 2
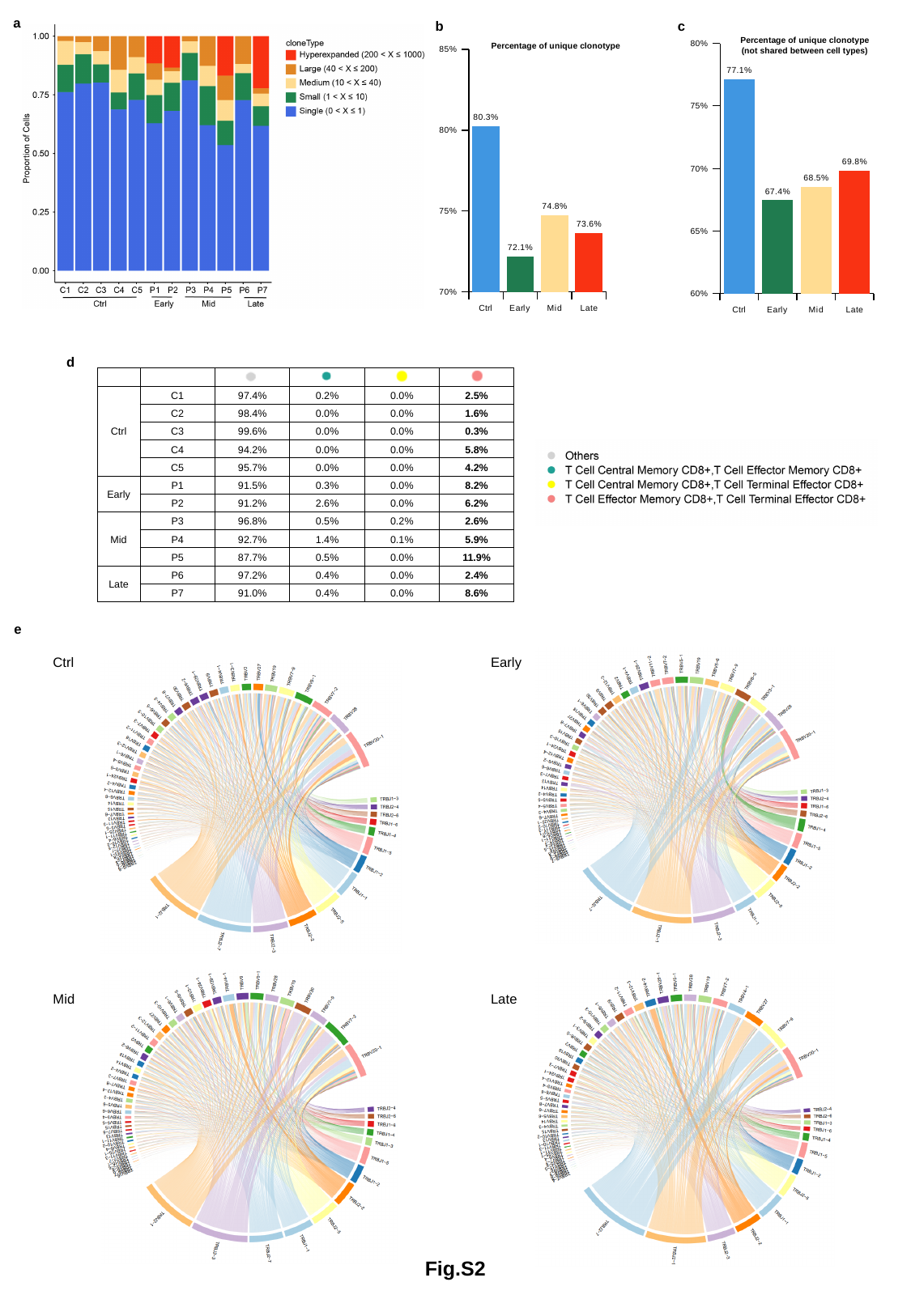

a
b
c
#### Chart
| Category | |
|---|---|
| Ctrl | 0.8026 |
| Early | 0.7216 |
| Mid | 0.7473 |
| Late | 0.7364 |Percentage of unique clonotype
Percentage of unique clonotype
(not shared between cell types)
#### Chart
| Category | |
|---|---|
| Ctrl | 0.7711845689464772 |
| Early | 0.6745643011732544 |
| Mid | 0.685319457476755 |
| Late | 0.6981760785689232 |d
| | | | | | |
| --- | --- | --- | --- | --- | --- |
| Ctrl | C1 | 97.4% | 0.2% | 0.0% | 2.5% |
| | C2 | 98.4% | 0.0% | 0.0% | 1.6% |
| | C3 | 99.6% | 0.0% | 0.0% | 0.3% |
| | C4 | 94.2% | 0.0% | 0.0% | 5.8% |
| | C5 | 95.7% | 0.0% | 0.0% | 4.2% |
| Early | P1 | 91.5% | 0.3% | 0.0% | 8.2% |
| | P2 | 91.2% | 2.6% | 0.0% | 6.2% |
| Mid | P3 | 96.8% | 0.5% | 0.2% | 2.6% |
| | P4 | 92.7% | 1.4% | 0.1% | 5.9% |
| | P5 | 87.7% | 0.5% | 0.0% | 11.9% |
| Late | P6 | 97.2% | 0.4% | 0.0% | 2.4% |
| | P7 | 91.0% | 0.4% | 0.0% | 8.6% |
e
Ctrl
Early
Mid
Late
Fig.S2

### Slide 3
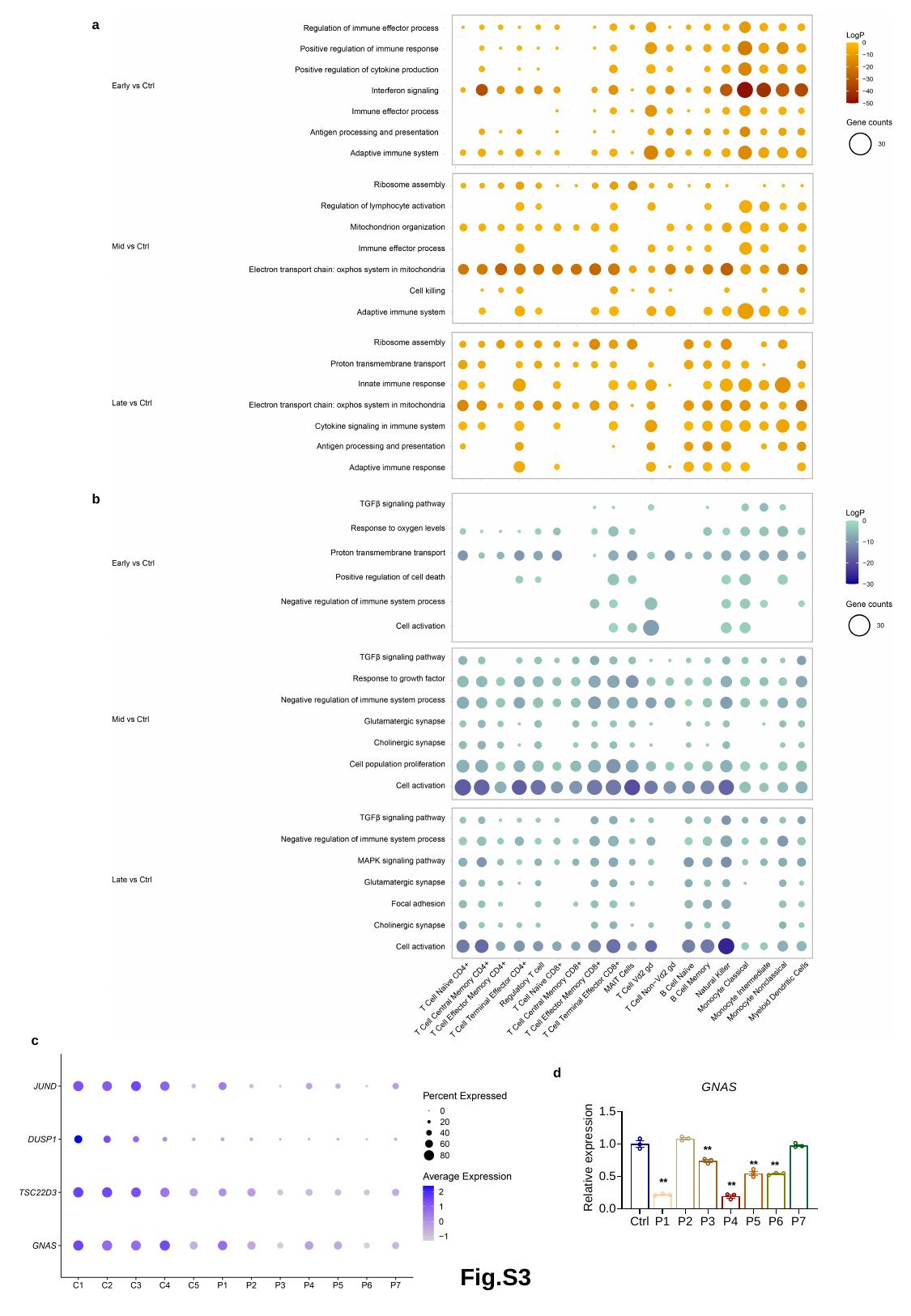

a
b
c
d
Fig.S3

### Slide 4
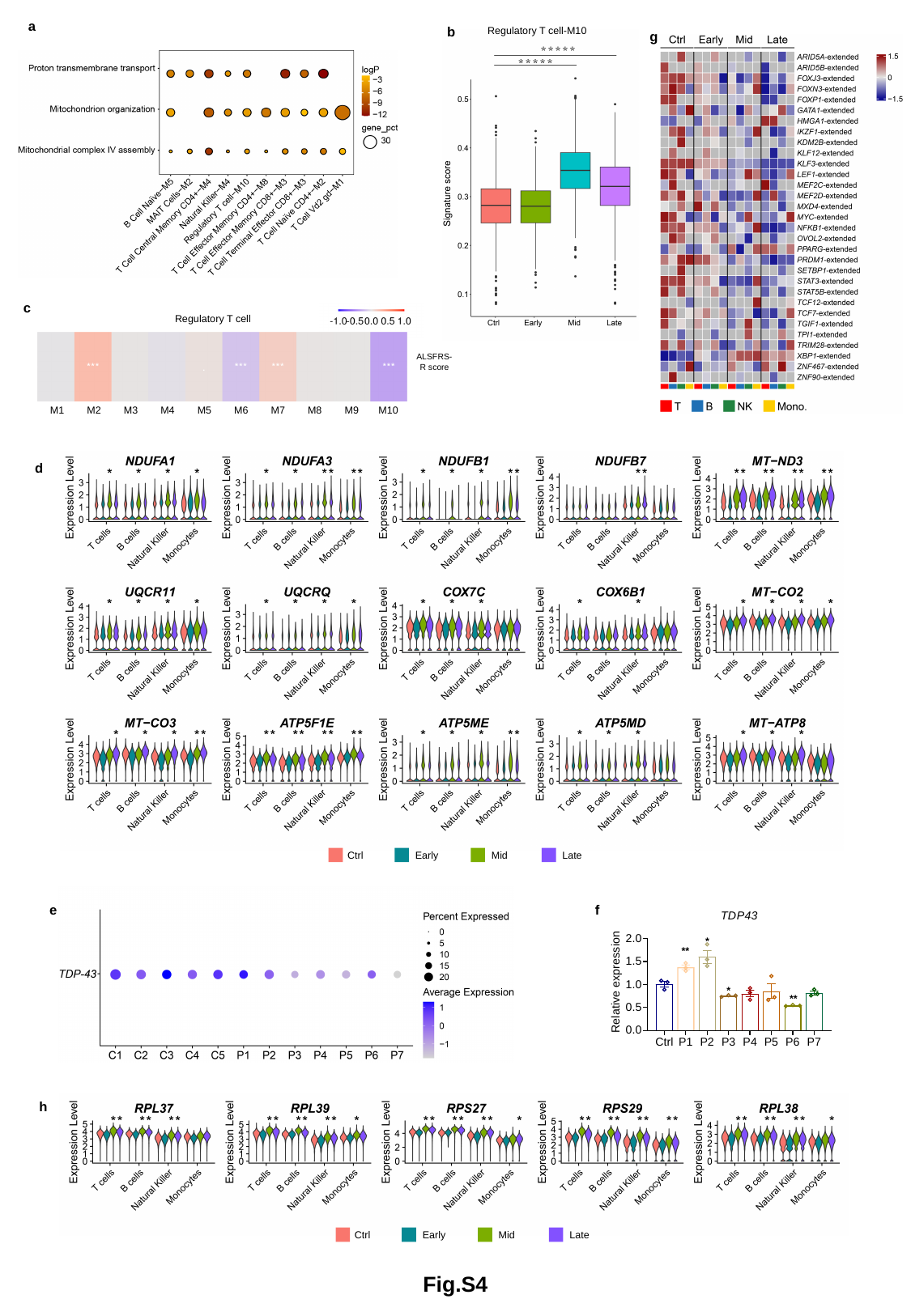

a
b
Regulatory T cell-M10
g
c
Regulatory T cell
ALSFRS-R score
M1
M2
M3
M4
M5
M6
M7
M8
M9
M10
d
Ctrl
Early
Mid
Late
e
f
h
Ctrl
Early
Mid
Late
Fig.S4

### Slide 5
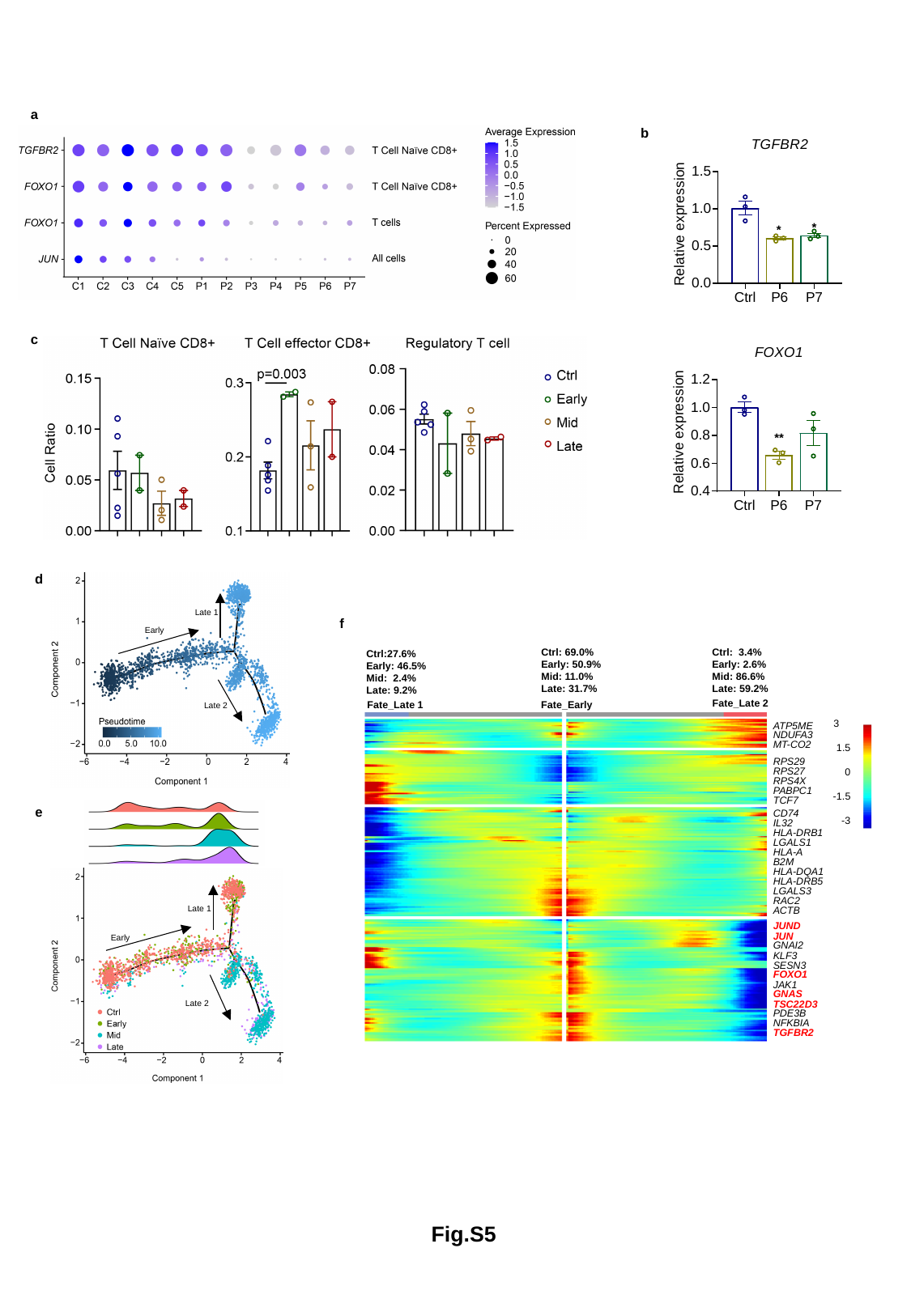

a
b
c
d
Late 1
Early
Late 2
f
Ctrl: 3.4%
Early: 2.6%
Mid: 86.6%
Late: 59.2%
Ctrl: 69.0%
Early: 50.9%
Mid: 11.0%
Late: 31.7%
Ctrl:27.6%
Early: 46.5%
Mid: 2.4%
Late: 9.2%
Fate_Late 2
Fate_Late 1
Fate_Early
3
1.5
0
-1.5
-3
ATP5ME
NDUFA3
MT-CO2
RPS29
RPS27
RPS4X
PABPC1
TCF7
CD74
IL32
HLA-DRB1
LGALS1
HLA-A
B2M
HLA-DQA1
HLA-DRB5
LGALS3
RAC2
ACTB
JUND
JUN
GNAI2
KLF3
SESN3
FOXO1
JAK1
GNAS
TSC22D3
PDE3B
NFKBIA
TGFBR2
e
Late 1
Early
Late 2
Fig.S5

### Slide 6
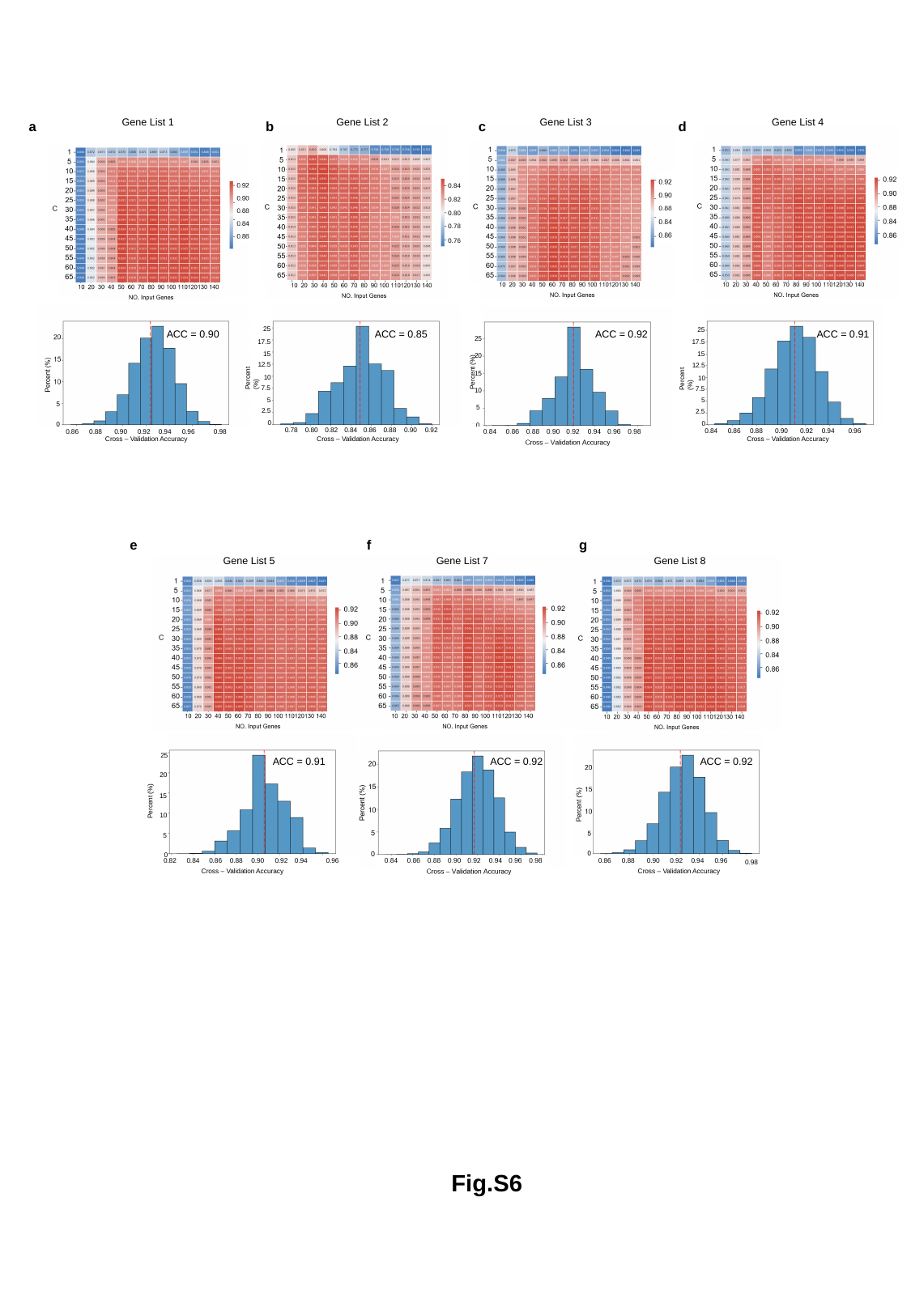

Gene List 1
Gene List 2
Gene List 3
Gene List 4
a
b
c
d
0.84
0.82
0.80
0.78
0.76
25
25
ACC = 0.90
ACC = 0.85
ACC = 0.92
ACC = 0.91
25
17.5
17.5
Percent (%)
15
15
20
12.5
12.5
Percent (%)
Percent (%)
15
10
10
7.5
7.5
10
5
5
5
2.5
2.5
0
0
0
0.78
0.80
0.82
0.84
0.86
0.88
0.90
0.92
0.84
0.86
0.88
0.90
0.92
0.94
0.96
0.86
0.88
0.90
0.92
0.94
0.96
0.98
Cross – Validation Accuracy
Cross – Validation Accuracy
Cross – Validation Accuracy
e
f
g
Gene List 5
Gene List 7
Gene List 8
25
ACC = 0.91
ACC = 0.92
ACC = 0.92
20
Percent (%)
15
10
5
0
0.82
0.84
0.86
0.88
0.90
0.92
0.94
0.86
0.88
0.90
0.92
0.94
0.96
0.96
0.98
Cross – Validation Accuracy
Cross – Validation Accuracy
Fig.S6
